# Examining Generalizability of Nutrient-Based Food Patterns and Their Cross-Sectional Associations with Cardiometabolic Health for Hispanic/Latino Adults in the US: Results from the National Health and Nutrition Examination Survey (NHANES) and the Hispanic Community Health Study/Study of Latinos (HCHS/SOL)

**DOI:** 10.1101/2023.05.04.23289531

**Authors:** Jeanette J Varela, Josiemer Mattei, Daniela Sotres-Alvarez, Yasmin Mossavar-Rahmani, Amanda C McClain, Luis E Maldonado, Martha L Daviglus, Briana J.K. Stephenson

**Affiliations:** Department of Biostatistics, Harvard T.H. Chan School of Public Health; Department of Nutrition, Harvard T.H. Chan School of Public Health; Department of Biostatistics, Gillings School of Global Public Health, University of North Carolina at Chapel Hill; Department of Epidemiology and Population Health, Albert Einstein College of Medicine; School of Exercise and Nutritional Sciences, College of Health and Human Services, San Diego State University College of Health and Human Services; Department of Population and Public Health Sciences, Keck School of Medicine, University of Southern California; Institute of Minority Health Research, University of Illinois at Chicago

## Abstract

**Background:** Ethnicity, cultural background, and geographic location differ significantly amongst the US Hispanic/Latino population. These characteristic differences can greatly define measured diet and its relationship with cardiometabolic disease, thus influencing generalizability of results.

**Objective:** We aimed to examine dietary patterns of Hispanic/Latino adults and their association with cardiometabolic risk factors (high cholesterol, hypertension, obesity, diabetes) across two representative studies with differing sampling strategies.

**Methods:** Data were collected from Mexican or Other Hispanic adult participants from 2007-2012 National Health and Nutrition Examination Survey (NHANES, n=3,209) and 2007-2011 Hispanic Community Health Survey/Study of Latinos (HCHS/SOL, n=13,059). Nutrient-based food patterns (NBFPs) were derived using factor analysis on nutrient intake data estimated from 24-hour dietary recalls and interpreted using common foods prominent in these nutrients. Cross-sectional association between NBFPs (quintiles) and cardiometabolic risk factors, defined by clinical measures and self-report, were estimated using survey-weighted logistic regression.

**Results:** Five NBFPs were identified in both studies: (1) meats, (2) grains/legumes, (3) fruits/vegetables, (4) dairy, and (5) fats/oils. Association to cardiometabolic risk factors differed by NBFP and study. In HCHS/SOL, persons in the highest quintile of meats NBFP had higher odds of diabetes (OR=1.43, 95%CI: 1.10, 1.86) and obesity (OR=1.36, 95%CI: 1.14, 1.63). Those in the lowest quintile of grains/legumes NBFP (OR=1.22, 95%CI: 1.02, 1.47) and the highest quintile of fats/oils (OR=1.26, 95%CI: 1.03, 1.53) also had higher odds of obesity. In NHANES, NBFPs associated with higher odds of diabetes included those in the lowest quintile of dairy (OR=1.66, 95%CI: 1.01, 2.72) and highest quintile of grains/legumes (OR=2.10, 95%CI: 1.26, 3.50). Persons in the fourth quintile of meats (OR=0.68, 95%CI: 0.47, 0.99) had lower odds of cholesterol.

**Conclusion:** Diet-disease relationships among Hispanic/Latino adults vary according to two representative studies. These differences have research and practical implications when generalizing inferences on heterogeneous underrepresented populations.

## 1 Introduction

Since 2010, Hispanic/Latino people have become the largest ethnic minority group in the US. Recent research found that Hispanics are nearly 10 years younger than non-Hispanic Whites at the time of death from cardiovascular disease (CVD) (1). Furthermore, due to limited access to healthy foods, higher prevalence of food insecurity, and lower socioeconomic status, US Hispanic/Latinos tend to consume less healthy diets when compared to other racial/ethnic groups (2), which then contributes to health complications including cardiovascular diseases (3). Thus, understanding the dietary patterns among Hispanics/Latinos in the US may help better address the CVD-related disparities experienced by this population.

Previous literature has shown that cardiometabolic risk factors (CRFs) differ among Hispanic/Latino ethnic groups due to diverse characteristics between groups such as place/country of birth, citizenship, language, race, culture, food, and other factors (4,5,6,7,8,9,10). Despite these known differences in Hispanic/Latino groups, nationally representative surveys continue to lump this population into two large groups; Mexican American and Other Hispanic/Latino. For example, the National Health and Nutrition Examination Survey (NHANES), which studies the health and nutritional status of people in the United States across varying socioeconomic status (SES), race/ethnicity, and geographies, aggregates Hispanic/Latinos into Mexican Americans and other Hispanics. Several studies have since followed suit reinforcing this limitation when evaluating the inter-relationship of diet and disease across different Hispanic/Latino groups (11,12,13,14,15,16).

The Hispanic Community Health Study/Study of Latinos (HCHS/SOL) was designed to better understand the health and well-being of this understudied population with data from six Hispanic/Latino ethnic backgrounds (Cuban, Dominican, Mexican, Puerto Rican, and Central/South American) across four US urban areas (Bronx, Chicago, Miami, San Diego). Both NHANES and HCHS/SOL contain vital information about Hispanic/Latino diets and CRFs, but little research has been done to compare how consistent these associations are across the two studies, given their differing sampling designs (17,18). Both diet quality and diet patterns have been examined within the HCHS/SOL cohort (19,20,21). Diet quality has been evaluated in NHANES Hispanic/Latino adults with children (22). While dietary patterns have been examined amongst Mexican American NHANES participants, to our knowledge, no studies have examined diet patterns exclusively for Mexican American and Other Hispanic adults. This is a significant gap as diet patterns provide a more comprehensive measure for examining diet-disease relationships (23).

This study aimed to derive and compare nutrient-based food patterns and their cross-sectional associations with cardiometabolic risk factors among US Hispanic/Latino adult participants from two different survey studies.

## 2 Methods

### 2.1 Study Population

This study included two U.S. studies that include Hispanic/Latino adults. Data were harmonized and only measures shared across both surveys were included for analysis. To minimize reverse causality, we excluded participants with previous CVD conditions (such as heart failure, coronary heart disease, angina, heart attack, or stroke) at time of enrollment.

We also excluded extreme nutrient intake defined as values below the 0.5th percentile and above the 99.5th percentile. The inclusion criteria for this analysis were adult participants aged 20-74 years old with at least one reliable recall that identify as Mexican American or other Hispanic groups (Supplementary Material Figure 1).

#### 2.1.1 National Health and Nutritional Examination Survey (NHANES)

The National Health and Nutrition Examination Survey (NHANES) program is a nationally representative repeated cross-sectional survey with a stratified, multistage probability sampling design of nonincarcerated residents of the United States. About 5,000 persons each year are interviewed and located in counties across the country. Details of the study design are described elsewhere (24). The data collected include demographic information, dietary intake, and health-related questions along with laboratory tests for three NHANES survey cycles (2007-08, 2009-10, and 2011-12). Dietary intake data were assessed via two 24-hour recalls which collects the types and amounts of foods and beverages consumed and allow estimated intakes of energy, nutrients, and other food components from those foods and beverages. The United States Department of Agriculture (USDA) Food and Nutrient Database for Dietary Studies 4.1 (25), 5.0 (26), and 2011-2012 (27) were used to code dietary intake data and calculate nutrient intakes for the NHANES 2007-08, 2009-10, and 2011-12 survey cycles respectively.

The first recall was collected in person. The second recall was administered over the telephone 3 to 10 days later. Nutrient intake data collected from the two interviews are publicly available on the CDC website (28,29,30). Due to data being pooled across three survey cycles (2007-2008, 2009-2010, 2011-2012), survey-weight adjustment was conducted (31,32). The inclusion criteria were adult participants (20-74 years old) that identify as Mexican American or other Hispanic groups as defined by NHANES, with at least one reliable recall (33). After excluding those with no prior CVD condition or extreme nutrient intake, a total of 1,930 Mexican American and 1,279 other Hispanic adults were included for analysis (Supplementary Material Figure 1A).

#### 2.1.2 Hispanic Community Health Survey/Study of Latinos (HCHS/SOL)

The Hispanic Community Health Study/Study of Latinos (HCHS/SOL) is a multi-center prospective study designed to identify risk factors and disease prevalence rates in a diverse population-based cohort of US Hispanic/Latino adults in four urban communities. From 2008-2011, HCHS/SOL recruited a cohort of 16,415 Hispanic/Latino persons aged 18–74 who self-identified as Cuban, Dominican, Mexican, Puerto Rican, and Central/South American and resided in households across four field centers in the US: Bronx, NY, Chicago IL, Miami FL, and San Diego CA. A stratified 2-stage area probability sample of household addresses was selected in each of the 4 field centers. Additional details on the design and sampling methods of HCHS/SOL have been previously described (34). Data on cardiometabolic risk factors, demographic information, and medical history were recorded by questionnaires. Dietary intake was assessed using two 24-hour dietary recalls and then used to calculate nutrient intake using the Nutrition Data System for Research software version 11 (35). The first recall was collected in person. The second recall was administered via telephone after 6 weeks. The inclusion criteria were adult participants aged 20-74 years old with at least one reliable recall according to the interviewer. After excluding those with no prior CVD condition or extreme nutrient intake, a total of 5,308 Mexican American and 7,751 other Hispanic adults were included for analysis (Supplementary Material Figure 1B).

### 2.2 Cardiometabolic Risk Factors (CRFs)

Our analysis focused on four major modifiable, manageable, or treatable cardiometabolic risk factors that were assessed in both surveys during the same collection years described previously: high cholesterol, obesity, diabetes, and hypertension. High cholesterol was defined as having total cholesterol greater than or equal to 240 mg/dL, LDL cholesterol greater than or equal to 160 mg/dL, HDL cholesterol less than 40 mg/dL, self-reported use of cholesterol-lowering medication, or self-reported physician diagnosis of hypercholesterolemia (36). Obesity was defined as a BMI of 30 kg/m^2^ or greater for participants 20-44 years of age and waist circumference (women greater than 88 cm, men greater than 102 cm) for participants 45-74 years of age (37,38,39). Diabetes was defined as having a fasting time greater than 8 hours and fasting plasma glucose of 126 mg/dL or greater, fasting time less than or equal to 8 hours and fasting glucose 200 mg/dL or greater, or post-OGTT glucose of 200 mg/dL or greater, an HbA1c of 6.5% or greater, self-reported diabetes medication use, or self-reported physician diagnosis (40). Hypertension (high blood pressure) was defined as having systolic blood pressure greater than 140 mm Hg, or diastolic blood pressure 90 mm Hg or greater, or self-reported hypertensive medication use (41).

The Framingham CVD 10-year risk score was also calculated in both surveys to estimate a 10-year risk for atherosclerotic cardiovascular disease event (e.g., coronary death, nonfatal myocardial infarction, stroke) (42). This measure is calculated using information on age, sex, total cholesterol, HDL cholesterol, systolic blood pressure, blood pressure-lowering medication use, diabetes status, and smoking status. For NHANES participants, the Framingham CVD risk score was created using the CVrisk R package (43). For HCHS/SOL participants, this score was derived by the HCHS/SOL Coordinating Center (44).

Sensitivity analysis was performed to determine the impact of including participants with CVD and/or cancer at baseline to account for potential change in diet due to these diagnoses.

### 2.3 Sociodemographic and behavioral variables

Covariates used in the analysis included age, energy intake, sex, Mexican vs other Hispanic heritage, educational attainment, annual household income, marital status, years living in the mainland U.S., employment status, self-reported smoking status, and self-reported alcohol use. NHANES classifies race/ethnic background as Mexican and Other Hispanic. In an effort to provide parallel analysis and comparisons, participants with a non-Mexican ethnic background in HCHS/SOL were combined.

### 2.4 Statistical Analysis

#### 2.4.1 Identification of Nutrient-Based Food Patterns (NBFPs)

A total of 39 nutrients were included for analysis (Table 2). Nutrient intake was averaged across 24hr dietary recalls, log-transformed via a log(1+x) transformation, and scaled to achieve normality. To identify nutrient-based food patterns, factor analysis that adjusts for survey sampling design (45,46) was performed separately on each study.

The number of factors to retain was determined by the following criteria: factor eigenvalue greater than 1, scree plot construction, and factor interpretability. We applied a varimax rotation to achieve a better-defined loading structure. Nutrients with a rotated factor loading greater than or equal to |0.60| were considered ‘dominant nutrients’ and considered in the description and clinical interpretation of that factor. Using Bartlett’s weighted least squares method, we computed factor scores that indicate the degree to which each subject’s diet conforms to one of the identified patterns. To assess the internal reproducibility of the identified patterns we calculated Cronbach’s coefficient alphas (47). Sensitivity analysis was performed to examine the consistency of the results with analysis using at least one recall versus the average of two recalls. To ease clinical interpretability, patterns were named using common foods that prominently contain the nutrients of each pattern and are referred to as nutrient-based food patterns (NBFPs). These names were reached upon the consensus of three co-authors with no unresolved disagreements.

#### 2.4.2 Association of NBFP Quintiles on CRFs

Quintile-based categories of factor scores were calculated for each survey separately, adjusting for survey design. We use the third quintile to define a moderate intake of each NBFP as the reference category. Survey-weighted logistic regressions were run for each CRF and each pattern as the primary exposure, and jointly with all derived factors included. Odds ratios (ORs) and corresponding 95% confidence intervals (CIs) were estimated after adjusting for age, energy intake, sex, Mexican vs other Hispanic heritage, educational attainment, household income, marital status, years living in the mainland U.S., employment status, self-reported smoking status, and self-reported alcohol use. We considered p-values less than 0.05 significant for all analyses. All analyses were performed in R version 3.6.3 (48) using psych (49), haven (50), survey (46), ggplot2 (51), tidyverse (52) packages.

## 3 Results

### 3.1 Descriptive analysis

Descriptive information of the two studies is provided in Table 1. Similar characteristics that were shared across both survey cohorts included approximately half were female, and most individuals were employed, had lived in the U.S. for more than 10 years, and were non-smokers. Differences between the two studies were also identified. More individuals in NHANES than the HCHS/SOL sample identified as Mexican, had attained an education level higher than high school diploma or GED, had higher household income, were more likely to be married, and reported currently using alcohol. A greater proportion of adults in NHANES were classified with high cholesterol levels while similar proportions of adults had diabetes, obesity, hypertension, and mean Framingham CVD 10-year risk scores in both studies. Table 2 shows the mean (SE), adjusted for age, of the 39 nutrients used in the factor analysis. Similar nutrient intake values were observed between the studies.

**Table 1:**
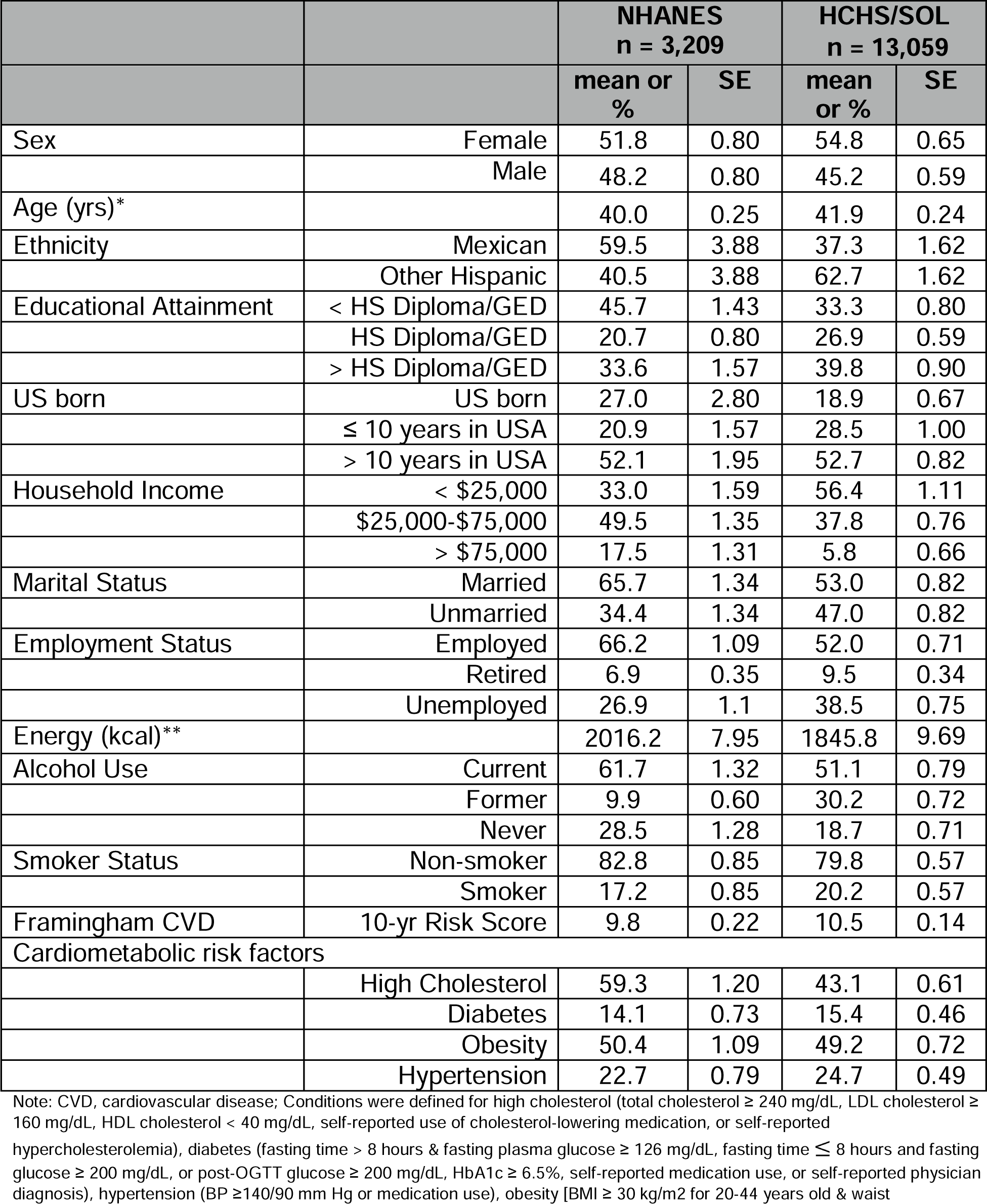
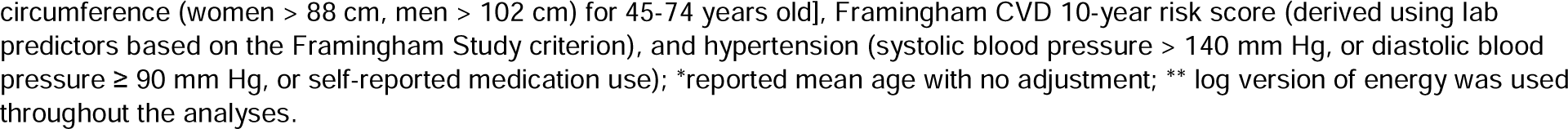
Sociodemographic and behavioral characteristics and cardiometabolic risk factors for NHANES (pooled cycles from 2008-2012) and HCHS/SOL (baseline 2008-2011) adjusting for age

|  |  | NHANES<br>n = 3,209 |  | HCHS/SOL<br>n = 13,059 |  |
| --- | --- | --- | --- | --- | --- |
|  |  | mean or % | SE | mean or % | SE |
| Sex | Female | 51.8 | 0.80 | 54.8 | 0.65 |
|  | Male | 48.2 | 0.80 | 45.2 | 0.59 |
| Age (yrs)* |  | 40.0 | 0.25 | 41.9 | 0.24 |
| Ethnicity | Mexican | 59.5 | 3.88 | 37.3 | 1.62 |
|  | Other Hispanic | 40.5 | 3.88 | 62.7 | 1.62 |
| Educational Attainment | < HS Diploma/GED | 45.7 | 1.43 | 33.3 | 0.80 |
|  | HS Diploma/GED | 20.7 | 0.80 | 26.9 | 0.59 |
|  | > HS Diploma/GED | 33.6 | 1.57 | 39.8 | 0.90 |
| US born | US born | 27.0 | 2.80 | 18.9 | 0.67 |
|  | ≤ 10 years in USA | 20.9 | 1.57 | 28.5 | 1.00 |
|  | > 10 years in USA | 52.1 | 1.95 | 52.7 | 0.82 |
| Household Income | < \$25,000 | 33.0 | 1.59 | 56.4 | 1.11 |
| | \$25,000-\$75,000 | 49.5 | 1.35 | 37.8 | 0.76 |
| | > \$75,000 | 17.5 | 1.31 | 5.8 | 0.66 |
| Marital Status | Married | 65.7 | 1.34 | 53.0 | 0.82 |
|  | Unmarried | 34.4 | 1.34 | 47.0 | 0.82 |
| Employment Status | Employed | 66.2 | 1.09 | 52.0 | 0.71 |
|  | Retired | 6.9 | 0.35 | 9.5 | 0.34 |
|  | Unemployed | 26.9 | 1.1 | 38.5 | 0.75 |
| Energy (kcal)** |  | 2016.2 | 7.95 | 1845.8 | 9.69 |
| Alcohol Use | Current | 61.7 | 1.32 | 51.1 | 0.79 |
|  | Former | 9.9 | 0.60 | 30.2 | 0.72 |
|  | Never | 28.5 | 1.28 | 18.7 | 0.71 |
| Smoker Status | Non-smoker | 82.8 | 0.85 | 79.8 | 0.57 |
|  | Smoker | 17.2 | 0.85 | 20.2 | 0.57 |
| Framingham CVD | 10-yr Risk Score | 9.8 | 0.22 | 10.5 | 0.14 |
| Cardiometabolic risk factors |  |  |  |  |  |
|  | High Cholesterol | 59.3 | 1.20 | 43.1 | 0.61 |
|  | Diabetes | 14.1 | 0.73 | 15.4 | 0.46 |
|  | Obesity | 50.4 | 1.09 | 49.2 | 0.72 |
|  | Hypertension | 22.7 | 0.79 | 24.7 | 0.49 |

Note: CVD, cardiovascular disease; Conditions were defined for high cholesterol (total cholesterol ≥ 240 mg/dL, LDL cholesterol ≥ 160 mg/dL, HDL cholesterol $<$ 40 mg/dL, self-reported use of cholesterol-lowering medication, or self-reported hypercholesterolemia), diabetes (fasting time $>$ 8 hours & fasting plasma glucose ≥ 126 mg/dL, fasting time ≤ 8 hours and fasting glucose ≥ 200 mg/dL, or post-OGTT glucose ≥ 200 mg/dL, HbA1c ≥ 6.5%, self-reported medication use, or self-reported physician diagnosis), hypertension (BP ≥ 140/90 mm Hg or medication use), obesity [BMI ≥ 30 kg/m<sup>2</sup> for 20-44 years old & waist
circumference (women > 88 cm, men > 102 cm) for 45-74 years old], Framingham CVD 10-year risk score (derived using lab predictors based on the Framingham Study criterion), and hypertension (systolic blood pressure > 140 mm Hg, or diastolic blood pressure ≥ 90 mm Hg, or self-reported medication use); \*reported mean age with no adjustment; \*\* log version of energy was used throughout the analyses.

### 3.2 Nutrient-Based Food Patterns

Five factors were retained for each study and accounted for 66.9% and 66.8% of the variance explained for NHANES and HCHS/SOL, respectively. Factors were similar between the two studies, but the proportion of variance explained between similar study-specific factors was different. A heatmap of the factor loadings is provided in Figure 1 and, to facilitate comparisons, both studies were ordered according to how the NHANES factors were retained.

**Figure 1:**
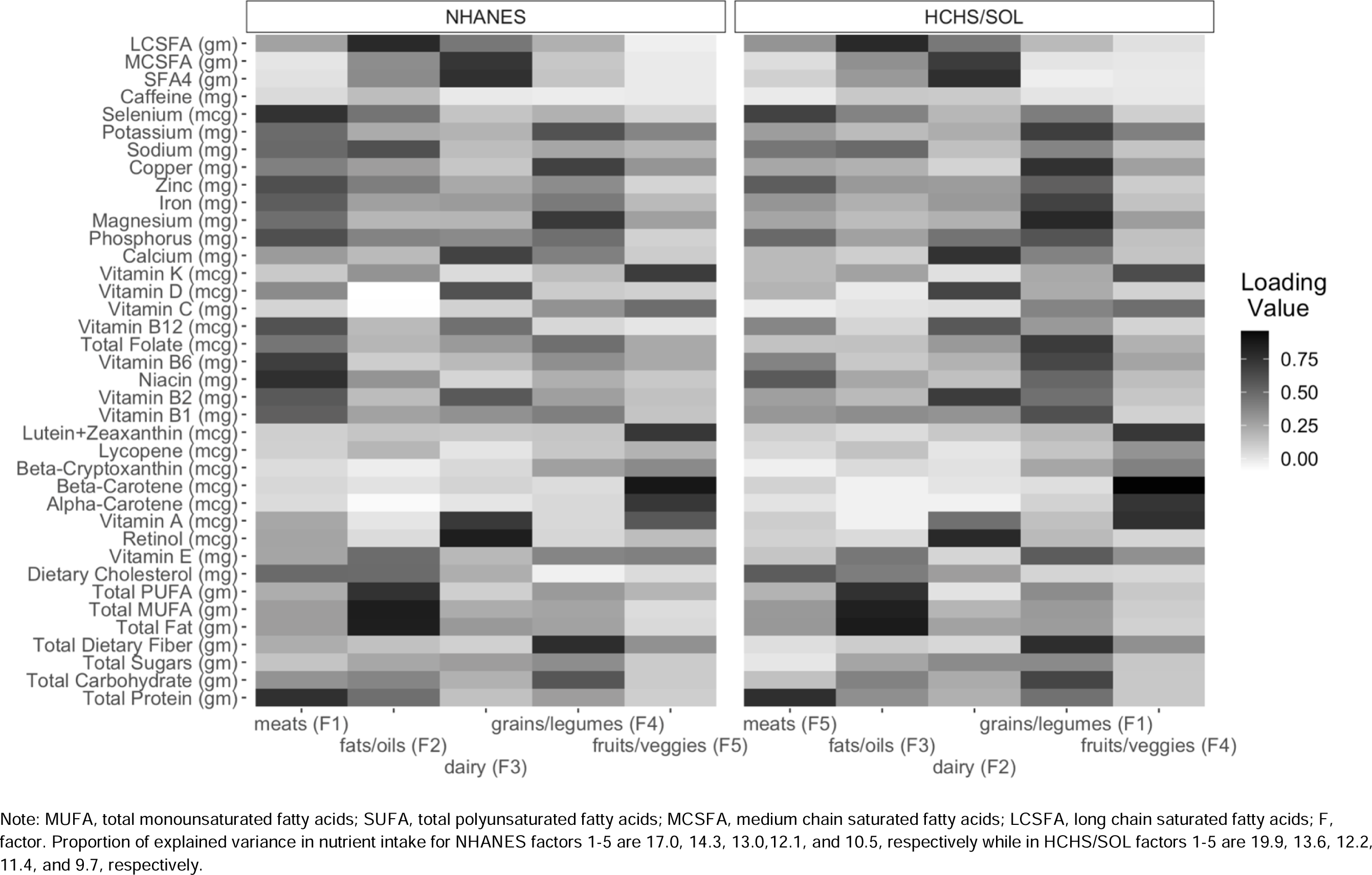
Heatmap of factor loading values for the five retained factors in NHANES and HCHS/SOL

NHANES Factor 1, named “meats”, had dominant loadings on total protein, niacin, vitamin B6, phosphorus, zinc, selenium, and vitamin B12. This factor alone explained 17.0% of the variance in nutrient intake. NHANES Factor 2, named “fats/oils”, had dominant loadings on total monounsaturated fatty acids (MUFA), total fat, long-chain saturated fatty acids (LCSFA), total polyunsaturated fatty acid (PUFA), and sodium. This factor explained 14.3% of the variance in nutrient intake. NHANES Factor 3, named “dairy”, had dominant loadings on retinol, vitamin A, calcium, butanoic (butyric acid or SFA 4:0), and minor-chain saturated fatty acids (MCSFA). The dairy factor explained 13.0% of the variance in nutrient intake. NHANES Factor 4, named “grains/legumes” had dominant loadings on copper, magnesium, and total dietary fiber. This factor explained 12.1% of the variance in nutrient intake. NHANES Factor 5, named “fruits/veggies”, loaded high on vitamin K, lutein + zeaxanthin, beta-carotene, and alpha-carotene. The fruits/veggies factor explained 10.5% of the variance in nutrient intake.

HCHS/SOL Factor 1, named “grains/legumes”, had dominant loadings on total dietary fiber, magnesium, copper, potassium, total carbohydrate, total folate, vitamin B1, vitamin B6, and iron. This factor explained 19.9% of the variance in nutrient intake. HCHS/SOL Factor 2, named “dairy”, had dominant loadings on retinol, vitamin B2, vitamin D, calcium, SFA4, and MCSFA. The dairy factor explained 13.6% of the variance in nutrient intake. HCHS/SOL Factor 3, named “fats/oils”, had dominant loadings on total fat, MUFA, PUFA, and LCSFA. This factor explained 12.2% of the variance in nutrient intake. HCHS/SOL Factor 4, named “fruits/veggies”, had the greatest loadings on vitamin A, alpha-carotene, beta-carotene, lutein + zeaxanthin, and vitamin K. The fruits/veggies factor explained 11.4% of the variance in nutrient intake. HCHS/SOL Factor 5, named “meats”, had dominant loadings on total protein and selenium. This factor explained 9.7% of the variance in nutrient intake.

**Table 2:** Nutrient intake summary (geometric mean and 95% CI) for NHANES and HCHS/SOL, adjusting for age, sex, and energy intake

|  | **NHANES**<br>**n = 3,209** | **HCHS/SOL**<br>**n = 13,059** |
| --- | --- | --- |
| **Nutrients** | **Geometric mean**<br>**(95% CI)** | **Geometric mean**<br>**(95% CI)** |
| Total Protein (gm) | 78.2 (77.0, 79.5) | 71.6 (70.7, 72.5) |
| Total Carbohydrate (gm) | 247.4 (244.2, 250.7) | 228.4 (226.3, 230.5) |
| Total Sugars (gm) | 99.3 (96.2, 102.5) | 88.7 (86.6, 90.8) |
| Total Dietary Fiber (gm) | 16.4 (15.8, 17.1) | 15.3 (14.9, 15.7) |
| Total Fat (gm) | 67.8 (66.5, 69.2) | 60.2 (59.4, 60.9) |
| Total MUFA (gm) | 24.6 (24.1, 25.2) | 21.9 (21.6, 22.3) |
| Total PUFA (gm) | 14.9 (14.5, 15.3) | 12.6 (12.4, 12.9) |
| Dietary Cholesterol (mg) | 250.0 (239.4, 261.1) | 216.1 (210.1, 222.3) |
| Vitamin E (mg) | 6.1 (5.9, 6.4) | 5.9 (5.8, 6.0) |
| Retinol (mcg) | 281.4 (266.1, 297.6) | 263.4 (252.0, 275.4) |
| Vitamin A (mcg) | 426.1 (406.4, 446.8) | 587.1 (567.0, 607.9) |
| Alpha-Carotene (mcg) | 102.9 (87.7, 120.9) | 120.1 (107.8, 133.7) |
| Beta-Carotene (mcg) | 947.3 (862.0, 1041.5) | 1110.6 (1044.3, 1181.3) |
| Beta-Cryptoxanthin (mcg) | 40.2 (35.2, 46.1) | 41.2 (37.6, 45.2) |
| Lycopene (mcg) | 1550.6 (1261.4, 1910.3) | 1028.2 (867.3, 1220.3) |
| Lutein+Zeaxanthin (mcg) | 708.9 (662.5, 758.7) | 613.9 (582.7, 647.0) |
| Vitamin B1 (mg) | 1.6 (1.5, 1.6) | 1.5 (1.5, 1.6) |
| Vitamin B2 (mg) | 1.9 (1.8, 1.9) | 1.7 (1.7, 1.7) |
| Niacin (mg) | 22.7 (22.1, 23.2) | 19.8 (19.5, 20.1) |
| Vitamin B6 (mg) | 1.9 (1.9, 2.0) | 1.8 (1.7, 1.8) |
| Total Folate (mcg) | 367.2 (356.5, 378.2) | 340.9 (334.3, 347.7) |
| Vitamin B12 (mcg) | 3.9 (3.8, 4.1) | 3.6 (3.5, 3.7) |
| Vitamin C (mg) | 62.3 (57.4, 67.7) | 64.0 (60.9, 67.2) |
| Vitamin D (mcg) | 3.5 (3.2, 3.7) | 3.8 (3.6, 3.9) |
| Vitamin K (mcg) | 61.0 (57.6, 64.6) | 53.0 (51.0, 55.0) |
| Calcium (mg) | 842.8 (816.1, 870.3) | 684.1 (667.5, 701.1) |
| Phosphorus (mg) | 1288.6 (1266.0, 1311.7) | 1086.9 (1072.7, 1101.4) |
| Magnesium (mg) | 278.5 (271.7, 285.5) | 255.9 (252.1, 259.8) |
| Iron (mg) | 13.8 (13.4, 14.1) | 12.4 (12.2, 12.6) |
| Zinc (mg) | 10.3 (10.1, 10.6) | 9.5 (9.4, 9.7) |
| Copper (mg) | 1.2 (1.2, 1.3) | 1.2 (1.2, 1.2) |
| Sodium (mg) | 3080.9 (3018.6, 3144.5) | 2839.4 (2791.8, 2887.8) |
| Potassium (mg) | 2486.6 (2431.3, 2543.2) | 2226.1 (2192.7, 2260.1) |
| Selenium (mcg) | 104.9 (102.7, 107.2) | 99.3 (97.9, 100.7) |
| Caffeine (mg) | 46.5 (39.0, 55.6) | 33.8 (30.0, 38.0) |
| SFA4 (butyric acid) (gm) | 0.5 (0.4, 0.5) | 0.4 (0.4, 0.4) |
| MCSFA (gm) | 1.2 (1.1, 1.3) | 1.1 (1.1, 1.2) |
| LCSFA (gm) | 19.2 (18.7, 19.7) | 17.1 (16.9, 17.4) |

Note: CI, confidence interval; MUFA, total monounsaturated fatty acids; SUFA, total polyunsaturated fatty acids; MCSFA, medium chain saturated fatty acids; LCSFA, long chain saturated fatty acids; gm, grams; mg, milligrams, mcg, micrograms

Standardized Cronbach’s coefficient alphas confirmed most of the nutrients contributed to high reliability and pattern characterization (see Supplementary Material Tables 1 and 2). Internal reproducibility of the two samples was also confirmed by NBFP commonly found in this population using the congruence coefficient (see Supplementary Material Table 3).

#### NBFPs Comparative Analysis

The meats factors of HCHS/SOL and NHANES had two similar dominant nutrients: total protein and selenium, however HCHS/SOL differed in that niacin, vitamin B6, phosphorus, zinc, and vitamin B12 were not included. The fats/oils factor also showed strong similarities across both studies, except for sodium (loading value of 0.61) only loading in NHANES. The dairy factor showed many similarities across both studies, but HCHS/SOL also included vitamin D, and B2 and NHANES included vitamin A as dominant nutrients. The grains/legumes factors were similar except for potassium, vitamin B1, vitamin B6, iron, total folate, and total carbohydrate loading only in HCHS/SOL and copper, magnesium, and total dietary fiber loading only in NHANES. Both fruit/vegetable factors reflected consumption of deep-colored fruits and vegetables, but vitamin A had a higher loading in HCHS/SOL, and not in NHANES.

The order of the retained factors differed for each study. In NHANES, meats was the first factor retained and explained most of the variation in nutrient intake (∼17%). In HCHS/SOL, meats was the last factor retained with 9.7% explained variation in nutrient intake. In NHANES, the second retained factor was fats/oils explaining 14.3% of the variation in nutrient intake while in HCHS/SOL fats/oils was the third retained factor with 12.2% of explained variance. In NHANES, the third retained factor and HCHS/SOL second retained factor were both dairy with similar values of explained variation (∼13%). The fourth retained factor in NHANES was grains/legumes with 12.1% which differed from HCHS/SOL. In HCHS/SOL, grains/legumes was the first factor retained which explained most of the variation in nutrient intake (∼20%). The last factor retained in NHANES was fruits/veggies which explained 10.5% of the variation of nutrient intake. This was lower than HCHS/SOL fourth retained factor, also fruits/veggies, that explaining 11.4% of variation in nutrient intake.

### 3.3 Quintile-based Factor Score Characteristics

Tables 3 and 4 describe sociodemographic, behavioral and cardiometabolic characteristics among those in the lowest and the highest quintile of each factor for NHANES and HCHS/SOL, respectively.

**Table 3:** Comparing NHANES characteristics by lowest and highest quintiles of factor scores

|  |  | Meats |  | Fats/Oils |  | Dairy |  | Grains/Legumes |  | Fruits/Veggies |  |
| --- | --- | --- | --- | --- | --- | --- | --- | --- | --- | --- | --- |
|  |  | Q1 | Q5 | Q1 | Q5 | Q1 | Q5 | Q1 | Q5 | Q1 | Q5 |
| Sex (%) | Female | 70.8 (3.9) | 20.3 (2.6) | 63.7 (3.7) | 33.9 (3) | 37.4 (3.3) | 43.1 (3.4) | 59.4 (3.7) | 33 (2.5) | 40.9 (3.5) | 51.3 (3.5) |
| Age (yrs) |  | 48.3 (0.8) | 43.4 (0.6) | 49.3 (0.8) | 43.8 (0.8) | 47.1 (0.9) | 45.6 (0.8) | 47.4 (0.9) | 45.0 (0.8) | 46.2 (0.9) | 47.6 (0.7) |
| Ethnicity (%) | Mexican | 61.7 (4.8) | 62.8 (4.9) | 52.1 (4.1) | 64.1 (5.8) | 64.9 (4.3) | 57.5 (4.9) | 52.1 (5.5) | 72.7 (3.8) | 61.5 (5.9) | 49.1 (5) |
| Educational Attainment (%) | < HS Diploma/GED | 51.0 (4.5) | 44.3 (3.3) | 60.2 (3.5) | 36.4 (3.9) | 55.7 (3.9) | 39.6 (3.1) | 46.5 (4) | 52.2 (4.2) | 61.4 (3.7) | 36.3 (3.1) |
|  | HS Diploma/GED | 20.7 (3.2) | 22.9 (3.1) | 15.2 (2.1) | 21.3 (3.3) | 19 (2.7) | 25.7 (3.4) | 16.4 (2.2) | 16.7 (3) | 16 (2.5) | 22.5 (3) |
|  | > HS Diploma/GED | 28.3 (4.4) | 32.8 (4.1) | 24.6 (2.7) | 42.3 (4.3) | 25.3 (3.2) | 34.7 (2.8) | 37.1 (3.6) | 31.1 (2.9) | 22.6 (3.3) | 41.2 (3.3) |
| US born (%) | US born | 25.4 (3.8) | 26.3 (3.4) | 12.5 (2.2) | 42.8 (4.9) | 21.7 (3.4) | 30.9 (3.6) | 32.9 (4.4) | 17.5 (3.2) | 26.8 (3.9) | 25.2 (3.2) |
|  | <= 10 years in USA | 16.8 (2.7) | 23.3 (3.1) | 17.7 (2.4) | 14.1 (3.2) | 14.7 (2.9) | 10.8 (2.1) | 17.3 (2.7) | 19.5 (3.5) | 19.2 (3.2) | 19.1 (3) |
|  | > 10 years in USA | 57.8 (3.9) | 50.4 (3.4) | 69.7 (2.7) | 43.1 (4.1) | 63.6 (2.6) | 58.3 (4) | 49.7 (4.2) | 63 (4.7) | 54.1 (4.4) | 55.6 (3.1) |
| Household Income (%) | < \$25,000 | 42.3 (4.7) | 27.7 (2.8) | 40 (3.5) | 24.9 (3.4) | 33 (3.6) | 32.8 (2.7) | 33.2 (3.3) | 29.7 (4.2) | 39.6 (4.2) | 31.2 (2.9) |
| | \$25,000-\$75,000 | 41.4 (3.2) | 56.6 (3.2) | 47.3 (3.1) | 52.6 (3.7) | 52.1 (3.5) | 45.5 (3.2) | 49.7 (2.7) | 58.3 (4.1) | 46.8 (4.2) | 47.2 (2.6) |
| | > \$75,000 | 16.4 (4.1) | 15.7 (2.9) | 12.7 (1.7) | 22.5 (4) | 14.9 (2.8) | 21.6 (3) | 17.2 (3.2) | 12 (2.5) | 13.5 (2) | 21.6 (2.9) |
| Marital Status (%) | Married | 64.6 (4) | 77 (2.9) | 64.1 (3.9) | 77.3 (3.1) | 68.5 (2.9) | 70.4 (3.9) | 62.2 (3.2) | 76.2 (3.1) | 68.5 (3.8) | 65.9 (2.7) |
| Employment Status (%) | Employed | 65.3 (2.9) | 77.5 (2.6) | 54.7 (3.1) | 81.8 (3.4) | 73.7 (3.3) | 69.3 (3.1) | 60 (3.4) | 80.1 (2.3) | 68.5 (3.6) | 70.4 (2.5) |
|  | Retired | 5.6 (1) | 3.9 (0.8) | 8.3 (1.3) | 2 (0.7) | 5.8 (1.4) | 5.5 (0.8) | 6.7 (1.2) | 3.2 (0.8) | 4.1 (1.1) | 6.4 (1) |
|  | Unemployed | 29.1 (2.4) | 18.5 (2.6) | 36.9 (2.7) | 16.2 (3) | 20.5 (2.7) | 25.3 (2.9) | 33.3 (3.1) | 16.7 (2.2) | 27.5 (3.1) | 23.1 (2.2) |
| Energy (kcal) |  | 1583.8<br>(33.4) | 2449.4<br>(61.8) | 1434.1<br>(29.4) | 2682.7<br>(48.1) | 1838.9<br>(51.3) | 2273.3<br>(62.7) | 1516.1<br>(40.3) | 2505.1<br>(62.7) | 1803.9<br>(44.4) | 2063.3<br>(61.5) |
| Alcohol Use (%) | Current | 45.7 (3.8) | 74 (3.1) | 47.1 (4.2) | 72.1 (2.7) | 58.7 (3.7) | 58.6 (3.6) | 55.6 (3.6) | 62.2 (3.6) | 56.6 (4.2) | 59.5 (3.9) |
|  | Former | 13.8 (2.2) | 9.1 (1.9) | 8.7 (1.7) | 9.7 (2) | 9.2 (2.2) | 10 (2.6) | 9.4 (2) | 12.5 (2.2) | 9.8 (1.7) | 12.7 (2.5) |
|  | Never | 40.6 (3.6) | 16.9 (2.6) | 44.1 (3.7) | 18.1 (2.8) | 32 (4.1) | 31.4 (3.3) | 35 (3.5) | 25.3 (3.1) | 33.6 (4.1) | 27.8 (3) |
| Smoker Status (%) | Smoker | 13.9 (3) | 21.3 (2.6) | 11.8 (2.2) | 21.1 (2.8) | 20 (2.6) | 16.1 (2.3) | 16.1 (2.7) | 14.3 (2.9) | 21.6 (3.3) | 13 (2.1) |
| Framingham CVD 10-yr | Risk Score | 8.5 (0.6) | 8.6 (0.4) | 9.8 (0.7) | 8.3 (0.7) | 10.9 (0.6) | 9.3 (0.8) | 8.5 (0.5) | 9.2 (0.5) | 10.3 (0.8) | 9.2 (0.5) |
| Cardiometabolic risk factors |  |  |  |  |  |  |  |  |  |  |  |
| High Cholesterol (%) | Yes | 57.2 (3.2) | 60.8 (3.7) | 58.3 (3.3) | 63.1 (3.2) | 61.3 (3.3) | 64.4 (2.8) | 63.2 (4) | 58.9 (3.3) | 65.6 (3.5) | 59.7 (3) |
| Diabetes (%) | Yes | 23.8 (3.8) | 14.6 (2.2) | 23.3 (3) | 17.4 (2.5) | 22.7 (2.8) | 18.5 (3) | 16.3 (3) | 23.0 (2.6) | 19.1 (3.4) | 19.5 (2) |
| Obesity (%) | Yes | 55.6 (3.6) | 49.8 (4.3) | 59.5 (3.1) | 60 (4.7) | 52.9 (4.1) | 50.7 (3.5) | 62.4 (3.9) | 47.8 (3.7) | 46.6 (2.9) | 58.1 (3.4) |
| Hypertension (%) | Yes | 30.7 (3.2) | 22.5 (2.6) | 31.4 (3.3) | 26.3 (2.3) | 29.9 (2.7) | 24.8 (3.7) | 26.7 (3.4) | 26.6 (3.5) | 24.5 (3.3) | 28.4 (2.5) |

Note: Q, quintile; CVD, cardiovascular disease; Conditions were defined for high cholesterol (total cholesterol ≥ 240 mg/dL, LDL cholesterol ≥ 160 mg/dL, HDL cholesterol $<$ 40 mg/dL, self-reported use of cholesterol-lowering medication, or self-reported hypercholesterolemia), diabetes (fasting time $>$ 8 hours & fasting plasma glucose ≥ 126 mg/dL, fasting time ≤ 8 hours and fasting glucose ≥ 200 mg/dL, or post-OGTT glucose ≥ 200 mg/dL, HbA1c ≥ 6.5%, self-reported medication use, or self-reported physician diagnosis), hypertension (BP ≥ 140/90 mm Hg or medication use), obesity [BMI ≥ 30 kg/m<sup>2</sup> for 20-44 years old & waist circumference (women $>$ 88 cm, men $>$ 102 cm) for 45-74 years old], Framingham CVD 10-year risk score (derived using lab predictors based on the Framingham Study criterion), and hypertension (systolic blood pressure $>$ 140 mm Hg, or diastolic blood pressure ≥ 90 mm Hg, or self-reported medication use). Dominant nutrients loading onto meats (protein, niacin, vitamin B6, phosphorus, zinc, selenium, vitamin B12), fats/oils (total MUFA, total fat, LCSFA, total PUFA, sodium), dairy (retinol, vitamin A, calcium, SFA4, MCSFA), grains/legumes (copper, magnesium, total dietary fiber), fruits/veggies (vitamin K, lutein+zeaxanthin, beta-carotene, alpha-carotene). Reference levels are male, other Hispanic, $>$ HS Diploma/GED, US born, annual income $>$ \$75k, married, employed, never drinker, and non-smoker.

**Table 4:** Comparing HCHS/SOL characteristics by lowest and highest quintiles of factor scores

|  |  | Grains/Legumes |  | Dairy |  | Fats/Oils |  | Fruits/Veggies |  | Meats |  |
| --- | --- | --- | --- | --- | --- | --- | --- | --- | --- | --- | --- |
|  |  | Q1 | Q5 | Q1 | Q5 | Q1 | Q5 | Q1 | Q5 | Q1 | Q5 |
| Sex (%) | Female | 67.4 (1.7) | 38.7 (1.6) | 54.1 (1.6) | 53.7 (1.7) | 68.5 (1.4) | 43.2 (1.9) | 54.8 (1.7) | 57.1 (1.7) | 72.1 (1.4) | 32.3 (1.6) |
| Age (yrs) |  | 47.3 (0.4) | 46.5 (0.4) | 47.0 (0.4) | 46.6 (0.5) | 48.6 (0.4) | 45.4 (0.4) | 46.6 (0.5) | 47.7 (0.4) | 49.2 (0.5) | 45.4 (0.4) |
| Ethnicity (%) | Mexican | 27.9 (2) | 46.6 (2.4) | 31.2 (2.3) | 45.9 (2.6) | 41.2 (2.1) | 32.4 (2.3) | 30.2 (2.2) | 42.1 (2.5) | 41.8 (2.2) | 32 (2.3) |
| Educational Attainment (%) | < HS Diploma/GED | 37.6 (1.6) | 33.9 (1.7) | 41.6 (1.7) | 30.9 (1.9) | 38.2 (1.4) | 29.1 (1.6) | 39.4 (1.8) | 29.6 (1.8) | 36.7 (1.5) | 30.6 (1.6) |
|  | HS Diploma/GED | 24.9 (1.4) | 24.2 (1.4) | 26.1 (1.5) | 27.1 (1.7) | 26.6 (1.5) | 28.1 (1.8) | 28.5 (1.8) | 24.4 (1.3) | 24.9 (1.4) | 28.0 (1.7) |
|  | > HS Diploma/GED | 37.5 (1.8) | 41.9 (1.9) | 32.3 (1.5) | 42 (1.9) | 35.2 (1.7) | 42.8 (1.7) | 32.1 (1.8) | 45.9 (2.0) | 38.4 (1.7) | 41.4 (1.9) |
| US born (%) | US born | 17.4 (1.4) | 11.2 (1.2) | 10.4 (1.1) | 18.8 (1.6) | 9.6 (1.0) | 20.9 (1.9) | 21.2 (1.9) | 11.7 (1.4) | 14.1 (1.1) | 15.5 (1.5) |
|  | <= 10 years in USA | 21.7 (1.5) | 27.5 (1.7) | 25.3 (1.8) | 24.8 (1.7) | 23.2 (1.6) | 28.2 (1.8) | 20.3 (1.4) | 27.5 (1.9) | 24.1 (1.5) | 23.9 (1.8) |
|  | > 10 years in USA | 60.9 (1.7) | 61.2 (1.7) | 64.2 (1.8) | 56.4 (2.0) | 67.2 (1.6) | 51.0 (1.9) | 58.5 (1.9) | 60.8 (2.0) | 61.8 (1.7) | 60.6 (1.9) |
| Household Income (%) | < \$25,000 | 62.4 (1.8) | 53.6 (1.9) | 60.9 (1.9) | 53.5 (2.4) | 59.5 (1.9) | 56.0 (2.0) | 64.9 (2.0) | 50.3 (2.1) | 58.8 (1.7) | 51.9 (2.1) |
| | \$25,000-\$75,000 | 32.7 (1.6) | 38.0 (1.6) | 35.4 (1.8) | 37.8 (1.9) | 35.2 (1.4) | 37.4 (1.8) | 30.5 (1.8) | 40.3 (1.7) | 36.1 (1.6) | 41.5 (1.8) |
| | > \$75,000 | 4.9 (0.9) | 8.4 (1.7) | 3.7 (0.7) | 8.7 (1.9) | 5.3 (1.1) | 6.6 (1.0) | 4.6 (0.9) | 9.3 (1.7) | 5.1 (0.8) | 6.6 (1.4) |
| Marital Status (%) | Married | 53.5 (1.8) | 62.1 (1.7) | 57.2 (1.7) | 60.4 (2.2) | 58.3 (1.7) | 55.6 (1.9) | 54.3 (2.0) | 63.5 (1.8) | 53.2 (1.7) | 58.0 (2.0) |
| Employment Status (%) | Employed | 49.7 (1.6) | 59.6 (1.7) | 53.5 (1.7) | 53.0 (1.9) | 48.6 (1.5) | 57.9 (1.9) | 48.7 (1.8) | 57.6 (1.8) | 48.5 (1.7) | 61.3 (1.8) |
|  | Retired | 13.4 (1.1) | 7.9 (1.0) | 9.6 (1.2) | 8.7 (0.9) | 14.0 (1.2) | 6.1 (1.1) | 10.3 (1.0) | 9.4 (1.2) | 14.6 (1.4) | 7.9 (1.2) |
|  | Unemployed | 37.0 (1.7) | 32.5 (1.6) | 37.0 (1.6) | 38.3 (2.0) | 37.4 (1.4) | 36.0 (1.9) | 40.9 (1.9) | 33.0 (1.7) | 36.9 (1.5) | 30.8 (1.6) |
| Energy (kcal) |  | 1285.3<br>(14.7) | 2360.0<br>(24.4) | 1627.2<br>(21.3) | 2048.3<br>(24.0) | 1305.8<br>(13.8) | 2386.6<br>(26.2) | 1628.5<br>(22.6) | 1854.9<br>(23.0) | 1470.7<br>(16.5) | 2142.4<br>(26.4) |
| Alcohol Use (%) | Current | 45.4 (1.6) | 56.4 (1.7) | 49.5 (1.8) | 50.4 (2.2) | 40.3 (1.6) | 56.6 (1.9) | 47.0 (1.8) | 49.1 (1.7) | 42.1 (1.5) | 60.0 (2.1) |
|  | Former | 34.0 (1.8) | 29.8 (1.6) | 31.6 (1.9) | 32.9 (1.8) | 38.7 (1.6) | 27.6 (1.7) | 34.5 (1.7) | 32.4 (1.7) | 36.7 (1.5) | 28.3 (1.9) |
|  | Never | 20.6 (1.5) | 13.8 (1.1) | 19.0 (1.4) | 16.7 (1.4) | 20.9 (1.4) | 15.8 (1.4) | 18.5 (1.4) | 18.4 (1.3) | 21.2 (1.3) | 11.7 (1.1) |
| Smoker Status (%) | Smoker | 20.4 (1.4) | 20.9 (1.3) | 21.7 (1.4) | 18.2 (1.5) | 11.1 (0.8) | 26.4 (1.7) | 25.8 (1.8) | 15.7 (1.1) | 15.4 (1.0) | 25.6 (1.5) |
| Framingham CVD 10-yr | Risk Score | 10.0 (0.4) | 10.0 (0.4) | 9.2 (0.4) | 10.1 (0.4) | 9.3 (0.3) | 10.0 (0.4) | 9.5 (0.4) | 9.7 (0.4) | 10.3 (0.4) | 10.0 (0.4) |
| Cardiometabolic risk factors |  |  |  |  |  |  |  |  |  |  |  |
| High Cholesterol (%) | Yes | 44.9 (1.6) | 48.2 (1.6) | 48.3 (1.6) | 45.0 (1.9) | 49.5 (1.5) | 43.5 (1.8) | 45.3 (1.9) | 45.2 (1.7) | 44.2 (1.6) | 49.7 (1.6) |
| Diabetes (%) | Yes | 18.8 (1.2) | 14.7 (1.0) | 19.2 (1.4) | 15.0 (1.2) | 22.2 (1.2) | 13.7 (1.3) | 16.4 (1.2) | 17.7 (1.3) | 17.1 (1.2) | 18.6 (1.5) |
| Obesity (%) | Yes | 59.7 (1.6) | 45.0 (1.6) | 53.5 (1.7) | 48.1 (1.9) | 53.5 (1.7) | 53.1 (1.8) | 54.7 (1.8) | 51.5 (1.8) | 55.5 (1.6) | 50.4 (1.9) |
| Hypertension (%) | Yes | 34.1 (1.6) | 26.1 (1.5) | 31.9 (1.6) | 26.1 (1.4) | 32.2 (1.5) | 27.0 (1.5) | 30.1 (1.6) | 28.6 (1.5) | 32.1 (1.5) | 28.9 (1.5) |

In NHANES, those in the highest quintile of meats were more likely to be male, married, have an annual household income of $25,000 – $75,000, current alcohol drinker, smoker, or have diabetes. Those in the highest quintile of fats/oils were more likely to be male, be of Mexican heritage, have an educational attainment greater than HS/GED diploma, currently drink alcohol, or be employed. Those in the lowest quintile of dairy were mostly composed of individuals with an educational attainment less than HS/GED diploma. Those in the lowest quintile of grains/legumes were more likely to be females, be of Mexican heritage, have lived in the USA for more than 10 years, or have obesity. Those in the lowest quintile of fruits/veggies were more likely to be of Mexican heritage, have an educational attainment less than a HS/GED diploma, be a smoker, or have obesity.

In HCHS/SOL, those in the highest quintile of meats were more likely to be male, other Hispanic heritage, employed, or a current drinker. Those in the lowest quintile of grains/legumes were more likely to be female, be other Hispanic heritage, have obesity, or not have hypertension. Those in the highest quintile of dairy were more likely to have an educational attainment greater than HS diploma/GED. Those in the lowest quintile of fruits/veggies were more likely to have an educational attainment less than HS diploma/GED. Those in the lowest quintile of fats/oils were more likely to be female or have an educational attainment level less than HS diploma/GED.

### 3.4 NBFPs Association to Cardiometabolic Risk Factors

The forest plots in Figure 2 show the ORs and the 95% CIs for all cardiometabolic risk factors by quintiles of the retained NBFPs scores, adjusted for confounders and comorbidities. An additional heatmap plot for the single factor model with ORs is provided in Supplementary Figure 2. Table 5 gives the ORs (95% CI) for all CRFs by quintiles of factor scores, and all the confounders in section 2.4 and comorbidities.

**Figure 2:**
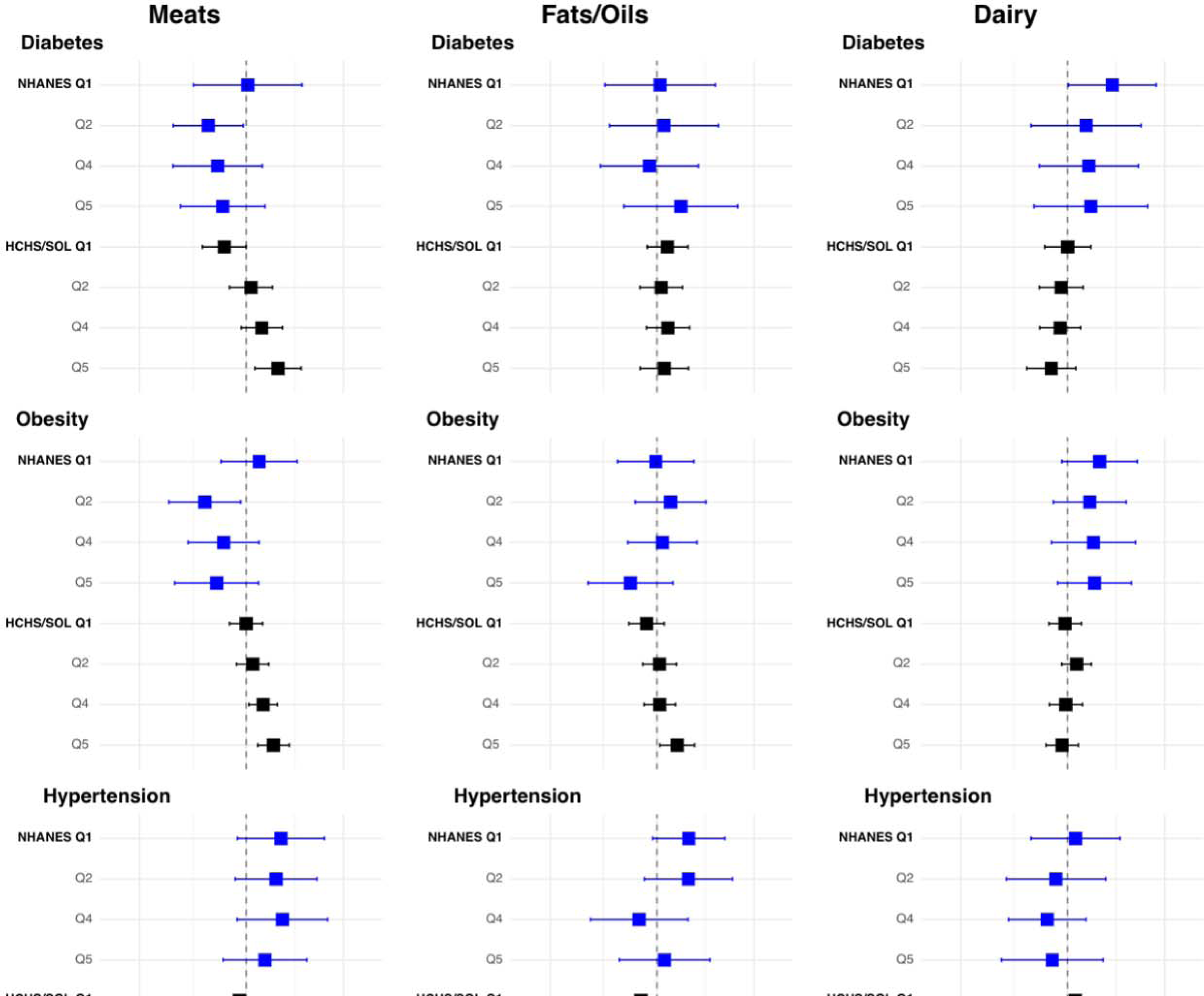
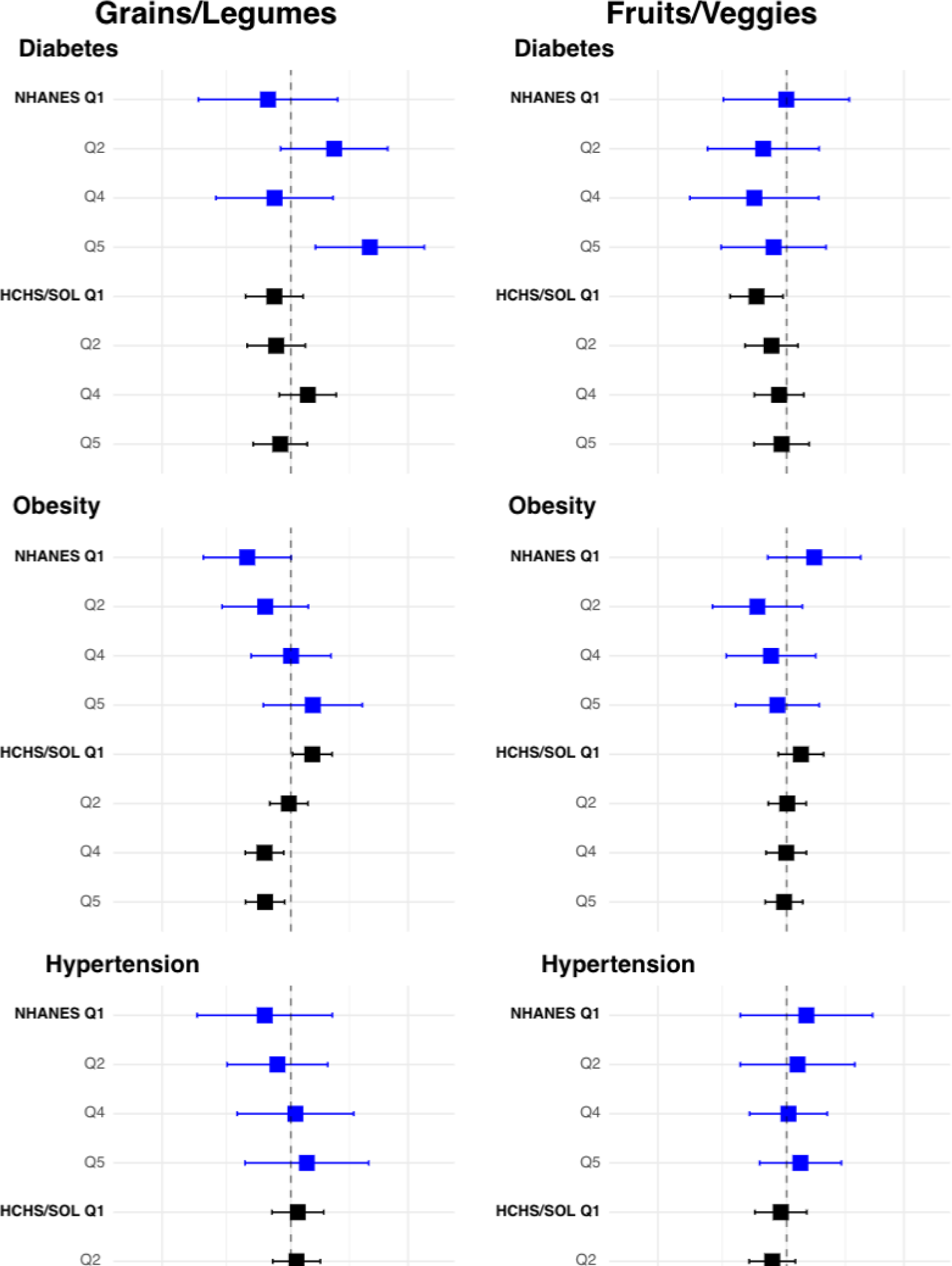
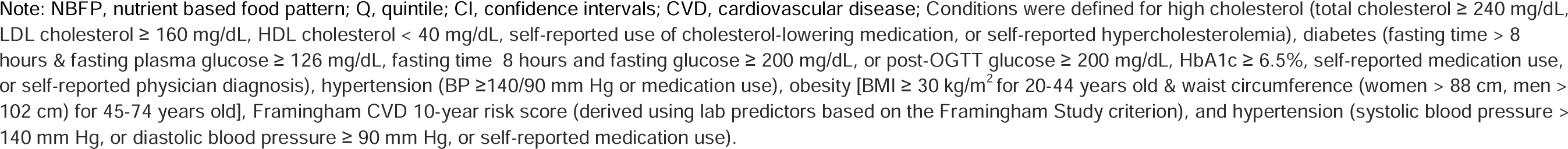
Forest plots of single factor models (OR and 95% CI) by NBFPs and cardiometabolic risk factors for NHANES (n=3,209) and HCHS/SOL (n=13,059) respondents

**Table 5:**
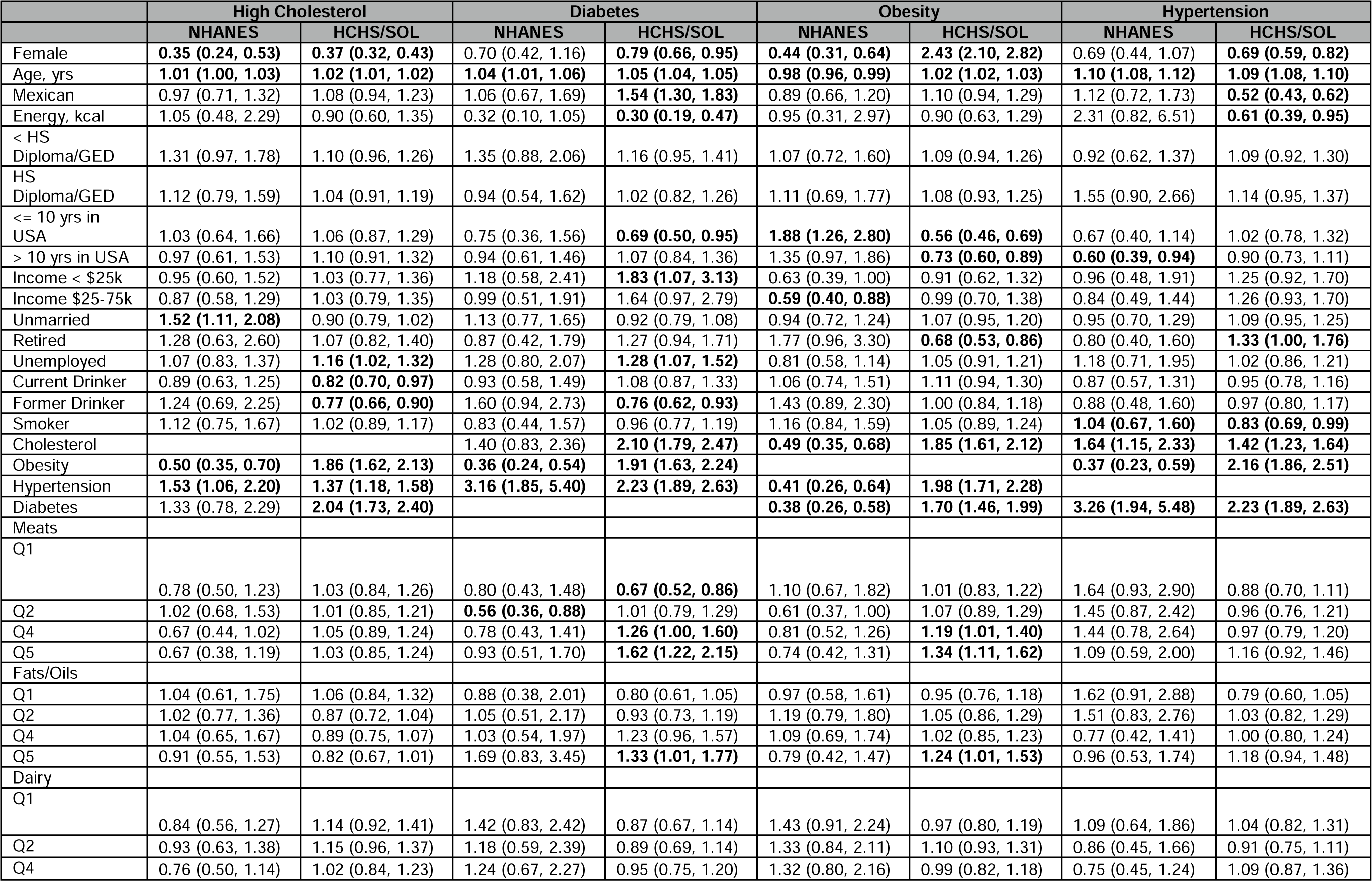
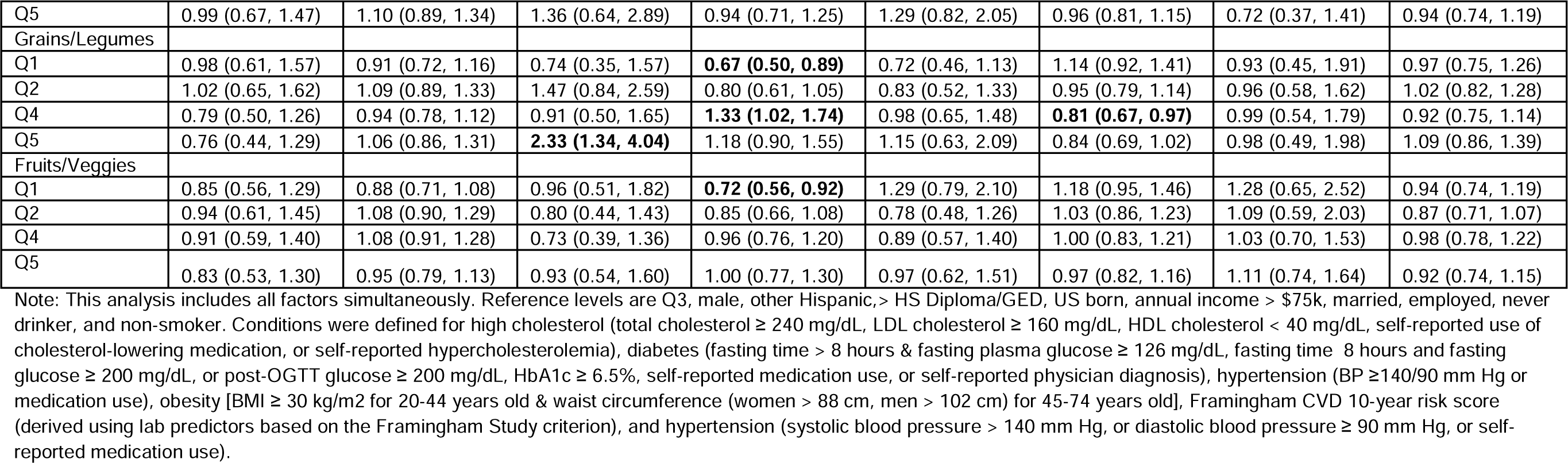
Odds ratio (95% CIs) for association of NHANES and HCHS/SOL respondents characteristics and dietary factors with cardiometabolic risk factors

#### Meats

For the meats NBFP, there were associations between meats and both diabetes and obesity were significant in both studies. In HCHS/SOL, persons in the highest quintile of meats had higher odds of diabetes and obesity, using the third quintile to define a moderate intake of meats for reference. This is similar to findings in NHANES where those in the second quintile of meats had lower odds of diabetes and obesity. In NHANES, the odds of cholesterol were lower for those in the fourth quintile of meats compared to the third quintile. No significant association was found between meats NBFP and hypertension in either study.

#### Fats/Oils

For the fats/oils NFBP, only one significant association emerged between the highest quintile of fats/oils and obesity in HCHS/SOL. There were no associations between fats/oils and any of the CRFs in NHANES. Compared to those in the third quintile of fats/oils, the odds of obesity were higher for persons in the highest quintile of fats/oils. No other associations were found between fats/oils NFBP and diabetes or high cholesterol in HCHS/SOL.

#### Fruits/Veggies

For the fruits/veggies NBFP, only one significant association was found for those with diabetes in HCHS/SOL. Compared to those in the third quintile of fruits/veggies, the odds of diabetes were lower for persons in the lowest quintile of fruits/veggies. No association was found between fruits/veggies NBFP and obesity, hypertension, or high cholesterol in either study.

#### Dairy

For the dairy NFBP, dairy was associated with diabetes in NHANES. Compared to those in the third quintile, higher odds of diabetes were observed among those in the lowest quintile of dairy in NHANES. No significant association was found between dairy NBFP and obesity, hypertension, or high cholesterol in either study.

#### Grains/Legumes

The odds of diabetes with intake of grains/legumes had fluctuating trends in NHANES. In NHANES, the odds of diabetes were significantly higher for those in the highest quintile of grains/legumes compared to those in the third quintile. In HCHS/SOL, the odds of obesity were lower for those in the highest quintile of grains/legumes when compared to those in the third quintile. No significant association was found between grains/legumes NBFP and hypertension or high cholesterol in either study.

## 4 Discussion

Our analysis identified five similar nutrient-based food patterns in HCHS/SOL and NHANES which were meats, fats/oils, dairy, grains/legumes, and fruits/vegetables. These factors explained ∼70% of the total variance in the nutrient intake. The order (importance) in which retained factors emerged differed between studies, which also indicates nutrient intake differences between studies. For example, the first factor retained in NHANES was meats, while in HCHS/SOL grains/legumes was retained first. When looking at characteristics by quintiles, we saw differences in intake patterns by study. For example, those in the highest quintile of meats in NHANES were more likely to be male, married, have an annual household income of $25,000 – $75,000, be a current alcohol drinker or smoker, or have diabetes, while those in HCHS/SOL that were in the highest quintile of meats were more likely to be male, other Hispanic heritage, employed, or current alcohol drinker. Despite these NBFPs similarities across studies, the associations between nutrient-based food patterns and cardiometabolic risk factors were distinctively different. Within the HCHS/SOL cohort, those in the highest quintile of meats were associated with higher odds of diabetes and obesity. NBFPs associated with higher odds of obesity included those in the highest quintile of fats/oils and meats and those in the lowest quintile of grains/legumes. There were no patterns associated with hypertension and high cholesterol in HCHS/SOL. Within the NHANES cohort, patterns associated with higher odds of diabetes included those in the lowest quintile of dairy and highest quintile of grains/legumes, with lower odds of obesity for those in the lowest quintile of meats and lower odds of cholesterol for those in the highest quintile of meats.

These discrepancies between studies raises concern of how best to generalize these differing results in characterizing dietary behaviors of US Hispanic/Latino adults (17,18). Each study has limitations regarding the population they represent partly explained by the study sampling design. This is further highlighted in Table 5, where additional demographic covariates included in the model yielded varying odds ratios between the two studies. HCHS/SOL implemented a sampling design aimed to collect an interpretable sized distribution of 7 identifiably different ethnic backgrounds in four large urban areas, omitting populations in non-urban areas. NHANES implemented a sampling design that aimed to collect an interpretable sized distribution relative to the US population for those that identify as Mexican or Other Hispanic in both urban or non-urban areas, but not publicly identifiable information to differentiate geography region or specify ethnicities of Other Hispanic adults (53). Further, while ample information about nativity and acculturation of participants was available for HCHS/SOL participants, this was largely unknown in NHANES. Differences in nativity and acculturation could potentially explain differences in the nutrients contained in the different foods consumed from the two studies, which could impact risks such as diabetes (54). These challenges limit the information we can obtain regarding nutrition and cardiometabolic health of this population from a single study in isolation.

Methods such as factor analysis, implemented in this study, or adherence scores to examine diet quality (14,55) rely on the composition of the study population. Greater representation of demographics that are known to influence diet (e.g., cultural background, geographical location) can drive the overall patterns identified in data-driven methods such as principal component analysis. Both studies include different Hispanic/Latino backgrounds, but the representation of Hispanic/Latino backgrounds vary between the two studies. With nutrient intake previously reported to differ by ethnic background in HCHS/SOL (4), different population compositions can yield different patterns between the two studies.

Prior work examining diets of Hispanic/Latino adult participants of NHANES or HCHS/SOL have focused on diet quality. Prior HCHS/SOL studies have analyzed adherence to diet quality scores and their associations to CVD by ethnic background (16, 21,55,56). Those of Mexican background had higher adherence to HEI-2015 or DASH with lower risk of cardiometabolic risk (e.g., blood pressure, fasting glucose, cholesterol) (55,56). This is consistent with our present HCHS/SOL findings, wherein persons in the highest quintile of meats NBFP had higher odds of diabetes and obesity compared to those in the third quintile. Nicklas et al. (14) examined diet quality (HEI-2005) differences by race/ethnicity of NHANES adult participants and its association with cardiometabolic risk. Their findings showed that mean HEI scores were higher among Other Hispanics but did not detail which dietary components contribute predominantly to that score. While other NHANES studies have focused primarily on individual nutrients of interest for Mexican American and Other Hispanic adults, none have focused primarily on deriving patterns of multiple nutrients in Hispanic/Latino adults (57,58). Given the unknown distribution of other Hispanic/Latino ethnic groups in NHANES, it is difficult to know how representative these results are over the growing diverse makeup of Hispanic/Latino adults in the United States. Advanced methods have been applied to HCHS/SOL to examine dietary differences by ethnicity and state (10,59), but similar approaches have not yet been explored with NHANES Hispanic/Latino participants for comparison.

We implemented an exploratory factor analysis in both studies to illustrate how a commonly used approach for deriving NBFP can impact generalized results of study populations, with similar ethnic backgrounds, but different survey sampling strategies. A strength of this study is the use of two 24hr dietary recalls from two large studies such as NHANES and HCHS/SOL. Due to NHANES oversampling Hispanic/Latino adults we had a moderately large sample size, allowing us enough power to analyze these two samples separately and compare results. The differences in sampling strategies permitted us to examine how different sampled populations can sometimes yield conflicting results. However, the different survey designs prevented us from being able to combine the two surveys and perform a direct comparison, such as a two-group CFA. Further methodological extensions are needed to allow pooling across multiple surveys with different sampling designs.

A limitation in our study is the use of 24-hour intake recall data which may affect the derivation of nutrient patterns when they fail to capture the participant’s usual diet and consumed nutrients, but this is ameliorated to a certain extent with the average of two recalls. While diet instruments are prone to underestimating energy intake, the 24-hour recalls have the strength given their granularity of capturing cultural and ethnic dietary differences versus food frequency questionaries (60). Thus, dietary recalls can be useful tools for studies of diverse ethnic composition. We would also like to acknowledge that self-reported dietary assessment tools and cardiometabolic risk factors are prone to measurement error and reporting bias (61,62). Another limitation is that nutrient patterns can differ by sex however in our derivation of NBFPs we did not adjust for sex. While sex was not used to derive nutrient-based food patterns, it was adjusted for in the associations with CRFs. The results reported in this study are based on a cross-sectional study design. Consequently, associations found are fixed at the time points listed. Changes in dietary behaviors or incidence of cardiometabolic risk factors over time are beyond the scope of this study.

In conclusion, this study demonstrates the sensitivity of diet patterns and their relation to cardiometabolic risk factors in US Hispanic/Latino adults, when different sampling strategies are implemented. While no single study can address all sampling strategy limitations, further methodological research should be explored to leverage already existing surveys that target underrepresented populations and account for study design differences, in order to generate appropriate population-based inference. For example, as mentioned previously, pooling studies focused on Hispanic/Latino adults whose sampling strategies differ by geography, race, ethnicity, and income would allow us to better examine the true heterogeneity of diet behaviors in the United States for this emerging demographic. This strategy will greatly improve population health disparities research by providing a more comprehensive understanding of nutrition and CVD epidemiology in populations at greatest risk.

## Data Availability

Data reported from the National Health and Nutrition Examination Survey is available for public use at: https://wwwn.cdc.gov/nchs/nhanes/. Data reported from the Hispanic Community Health Study/Study of Latinos is available through a data use agreement, upon request at:https://sites.cscc.unc.edu/hchs/.

## Abbreviations

NHANES: National Health and Nutritional Examination Survey
HCHS/SOL: Hispanic Community Health Study/ Study of Latinos
CRF: cardiometabolic risk factor
CVD: cardiovascular disease
USDA: United States Department of Agriculture
LCSFA: long chain saturated fatty acids
SFA 14:0: myristic acid
SFA 16:0: palmitic acid
SFA 18:0: stearic acid
MCSFA: medium chain saturated fatty acids
SFA 8:0: caprylic acid
SFA 6:0: caproic acid
SFA 10:0: capric acid
SFA 12:0: lauric acid
ORs: odds ratios
CIs: confidence intervals
SE: standard error
MUFA: total monounsaturated fatty acids
PUFA: total polyunsaturated fatty acid
HEI: Healthy Eating Index
NBFP: nutrient-based food pattern
SES: socioeconomic status

## References

1. Manjunath L, Hu J, Palaniappan L, Rodriguez F. Years of Potential Life Lost from Cardiovascular Disease Among Hispanics. Ethn Dis. 2019;29(3):477–484. doi:10.18865/ed.29.3.477

2. Ayala GX, Baquero B, Klinger S. A systematic review of the relationship between acculturation and diet among Latinos in the United States: implications for future research. J Am Diet Assoc. 2008;108(8):1330–1344. doi:10.1016/j.jada.2008.05.009

3. Wartella EA, Lichtenstein AH, Boon CS, editors. Front-of-Package Nutrition Rating Systems and Symbols: Phase I Report. Washington (DC): National Academies Press (US); Institute of Medicine (US) Committee on Examination of Front-of-Package Nutrition Rating Systems and Symbols; 2010. 4, Overview of Health and Diet in America. https://www.ncbi.nlm.nih.gov/books/NBK209844/

4. Siega-Riz AM, Sotres-Alvarez D, Ayala GX, et al. Food-group and nutrient-density intakes by Hispanic and Latino backgrounds in the Hispanic Community Health Study/Study of Latinos. Am J Clin Nutr. 2014;99(6):1487–1498. doi:10.3945/ajcn.113.082685

5. Rodriguez CJ, Allison M, Daviglus ML, Isasi CR, Keller C, Leira EC, Palaniappan L, Piña IL, Ramirez SM, Rodriguez B, Sims M; on behalf of the American Heart Association Council on Epidemiology and Prevention, Council on Clinical Cardiology, and Council on Cardiovascular and Stroke Nursing. Status of cardiovascular disease and stroke in Hispanics/Latinos in the United States: a science advisory from the American Heart Association. Circulation. 2014;130:593–625.

6. Daviglus ML, Pirzada A, Talavera GA. Cardiovascular disease risk factors in the Hispanic/Latino population: lessons from the Hispanic Community Health Study/Study of Latinos (HCHS/SOL). Prog Cardiovasc Dis. 2014;57(3):230–236. doi:10.1016/j.pcad.2014.07.006

7. Elfassy T, Zeki Al, Hazzouri A, Cai J, et al. Incidence of Hypertension Among US Hispanics/Latinos: The Hispanic Community Health Study/Study of Latinos, 2008 to 2017. J Am Heart Assoc. 2020;9(12):e015031. doi:10.1161/JAHA.119.015031

8. Maldonado LE, Adair LS, Sotres-Alvarez D, Mattei J, Mossavar-Rahmani Y, Perreira KM, Daviglus ML, Van Horn LV, Gallo LC, Isasi CR, Albrecht SS. Dietary Patterns and Years Living in the United States by Hispanic/Latino Heritage in the Hispanic Community Health Study/Study of Latinos (HCHS/SOL), The Journal of Nutrition, Volume 151, Issue 9, September 2021, Pages 2749–2759, https://doi.org/10.1093/jn/nxab165

9. Stephenson BJK, Sotres-Alvarez D, Siega-Riz AM, Mossavar-Rahmani Y, Daviglus ML, Van Horn L, Herring AH, Cai J. Empirically Derived Dietary Patterns Using Robust Profile Clustering in the Hispanic Community Health Study/Study of Latinos, The Journal of Nutrition, Volume 150, Issue 10, October 2020, Pages 2825–2834, https://doi.org/10.1093/jn/nxaa208

10. De Vito R, Stephenson B, Sotres-Alvarez D, Siega-Riz AM, Mattei J, Parpinel M, Peters BA, Bainter SA, Daviglus ML, Van Horn L, Edefonti V. Shared and ethnic background site-specific dietary patterns in the Hispanic Community Health Study/Study of Latinos (HCHS/SOL), Preprint, medRxiv 2022.06.30.22277013; doi: https://doi.org/10.1101/2022.06.30.22277013

11. Krok-Schoen JL, Archdeacon Price A, Luo M. et al. Low Dietary Protein Intakes and Associated Dietary Patterns and Functional Limitations in an Aging Population: A NHANES Analysis. J Nutr Health Aging 23, 338–347 (2019). https://doi.org/10.1007/s12603-019-1174-1

12. Ganji V, Shi Z, Al-Abdi T, Al Hejap D, Attia Y, Koukach D, Elkassas H. (2022). Association between food intake patterns and serum vitamin D concentrations in US adults. British Journal of Nutrition, 1–11. doi:10.1017/S0007114522001702

13. Aqeel MM, Guo J, Lin L, Gelfand SB, Delp EJ, Bhadra A, Richards EA, Hennessy E, Eicher-Miller HA, Temporal Dietary Patterns Are Associated with Obesity in US Adults, The Journal of Nutrition, Volume 150, Issue 12, December 2020, Pages 3259–3268, https://doi.org/10.1093/jn/nxaa287

14. Nicklas TA, O’Neil CE, Fulgoni VL, III, Diet Quality Is Inversely Related to Cardiovascular Risk Factors in Adults, The Journal of Nutrition, Volume 142, Issue 12, December 2012, Pages 2112–2118, https://doi.org/10.3945/jn.112.164889

15. Ha K, Sakaki JR, Chun OK, Nutrient Adequacy Is Associated with Reduced Mortality in US Adults, The Journal of Nutrition, Volume 151, Issue 10, October 2021, Pages 3214–3222, https://doi.org/10.1093/jn/nxab240

16. Chen F, Du M, Blumberg JB, et al. Association Among Dietary Supplement Use, Nutrient Intake, and Mortality Among U.S. Adults: A Cohort Study. Annals of Internal Medicine. 2019 May;170(9):604–613. DOI: 10.7326/m18-2478. PMID: 30959527; PMCID: PMC6736694.

17. Edefonti V, De Vito R, Salvatori A, Bravi F, Patel L, Dalmartello M, Ferraroni M. Reproducibility of A Posteriori Dietary Patterns across Time and Studies: A Scoping Review. Adv Nutr. 2020 Sep 1;11(5):1255–1281. doi: 10.1093/advances/nmaa032. PMID: 32298420; PMCID: PMC7490165.

18. Edefonti V, De Vito R, Dalmartello M, Patel L, Salvatori A, Ferraroni M. Reproducibility and Validity of A Posteriori Dietary Patterns: A Systematic Review. Advances in Nutrition (Bethesda, Md.). 2020 Mar;11(2):293–326. DOI: 10.1093/advances/nmz097. PMID: 31578550; PMCID: PMC7442345.

19. Maldonado LE. (2020). A Posteriori Dietary Patterns, Years Living in the US, and Type 2 Diabetes by Cultural Heritage: Unpacking Heterogeneity among Hispanics/Latinos. ProQuest Dissertations Publishing.

20. Siega-Riz AM, Pace ND, Butera NM, Van Horn L, Daviglus ML, Harnack L, Mossavar-Rahmani Y, Rock CL, Pereira RI, & Sotres-Alvarez D. (2019). How Well Do U.S. Hispanics Adhere to the Dietary Guidelines for Americans? Results from the Hispanic Community Health Study/Study of Latinos. Health equity, 3(1), 319–327. https://doi.org/10.1089/heq.2018.0105

21. Mattei J, Sotres-Alvarez D, Daviglus ML, Gallo LC, Gellman M, Hu FB, Tucker KL, Willett WC, Siega-Riz AM, Van Horn L, Kaplan RC, Diet Quality and Its Association with Cardiometabolic Risk Factors Vary by Hispanic and Latino Ethnic Background in the Hispanic Community Health Study/Study of Latinos, The Journal of Nutrition, Volume 146, Issue 10, October 2016, Pages 2035–2044, https://doi.org/10.3945/jn.116.231209

22. Overcash F, Reicks M. Diet Quality and Eating Practices among Hispanic/Latino Men and Women: NHANES 2011–2016. International Journal of Environmental Research and Public Health. 2021; 18(3):1302. https://doi.org/10.3390/ijerph18031302

23. Hu FB, Rimm EB, Stampfer MJ, Ascherio A, Spiegelman D, Willett WC. Prospective study of major dietary patterns and risk of coronary heart disease in men. Am J Clin Nutr. 2000;72(4):912–921. PMID: 11010931 https://doi.org/10.1161/circ.129.suppl_1.p113

24. Centers for Disease Control and Prevention (CDC). National Center for Health Statistics (NCHS). National Health and Nutrition Examination Survey Data. Hyattsville, MD: U.S. Department of Health and Human Services, Centers for Disease Control and Prevention, 2022, https://wwwn.cdc.gov/nchs/nhanes/analyticguidelines.aspx#sample-designhttps://wwwn.cdc.gov/nchs/nhanes/analyticguidelines.aspx#sample-design

25. USDA Food and Nutrient Database for Dietary Studies, 4.1 (2010). Beltsville, MD: U.S. Department of Agriculture, Agricultural Research Service, Food Surveys Research Group

26. Ahuja JKA, Montville JB, Omolewa-Tomobi G, Heendeniya KY, Martin CL, Steinfeldt LC, Anand J, Adler ME, LaComb RP, and Moshfegh AJ. 2012. USDA Food and Nutrient Database for Dietary Studies, 5.0. U.S. Department of Agriculture, Agricultural Research Service, Food Surveys Research Group, Beltsville, MD.https://wwwn.cdc.gov/nchs/nhanes/analyticguidelines.aspx#sample-design

27. U.S. Department of Agriculture, Agricultural Research Service. 2014. USDA Food and Nutrient Database for Dietary Studies 2011-2012. Food Surveys Research Group Home Page, http://www.ars.usda.gov/ba/bhnrc/fsrg

28. Centers for Disease Control and Prevention, National Center for Health Statistics. National Health and Nutrition Examination Survey 2007-2008 Data Documentation, Codebook, and Frequencies (Dietary Interview - Total Nutrient Intakes, First and Second Day, DR1TOT_E and DR2TOT_E). Hyattsville, MD: U.S. Department of Health and Human Services, Centers for Disease Control and Prevention, 2007-08. https://wwwn.cdc.gov/Nchs/Nhanes/2007-2008/DR1TOT_E.htm, https://wwwn.cdc.gov/Nchs/Nhanes/2007-2008/DR2TOT_E.htm2TOT_E.htm

29. Centers for Disease Control and Prevention, National Center for Health Statistics. National Health and Nutrition Examination Survey 2009-2010 Data Documentation, Codebook, and Frequencies (Dietary Interview - Total Nutrient Intakes, First and Second Day, DR1TOT_F and DR2TOT_F). Hyattsville, MD: U.S. Department of Health and Human Services, Centers for Disease Control and Prevention, 2009-10. https://wwwn.cdc.gov/Nchs/Nhanes/2009-2010/DR1TOT_F.htm, https://wwwn.cdc.gov/Nchs/Nhanes/2009-2010/DR2TOT_F.htm

30. Centers for Disease Control and Prevention, National Center for Health Statistics. National Health and Nutrition Examination Survey 2011-2012 Data Documentation, Codebook, and Frequencies (Dietary Interview - Total Nutrient Intakes, First and Second Day, DR1TOT_G and DR2TOT_G). Hyattsville, MD: U.S. Department of Health and Human Services, Centers for Disease Control and Prevention, 2011-12. https://wwwn.cdc.gov/Nchs/Nhanes/2011-2012/DR1TOT_G.htm, https://wwwn.cdc.gov/Nchs/Nhanes/2011-2012/DR2TOT_G.htm

31. Mirel LB, Mohadjer LK, Dohrmann SM, et al. National Health and Nutrition Examination Survey: Estimation procedures, 2007–2010. National Center for Health Statistics. Vital Health Stat 2(159). 2013.

32. Chen TC, Parker JD, Clark J, Shin HC, Rammon JR, Burt VL. National Health and Nutrition Examination Survey: Estimation procedures, 2011–2014. National Center for Health Statistics. Vital Health Stat 2(177). 2018.https://wwwn.cdc.gov/nchs/nhanes/analyticguidelines.aspx

33. Anand J, Raper N, Tong A. (2006). Quality assurance during data processing of food and nutrient intakes. Subtropical plant science, 19, S86. doi:

34. Lavange LM, Kalsbeek WD, Sorlie PD, Aviles-Santa LM, Kaplan RC, Barnhart J, Liu K, Giachello A, Lee DJ, Ryan J, et al. Sample design and cohort selection in the Hispanic Community Health Study/Study of Latinos. Ann Epidemiol 2010;20:642–9.

35. Schakel SF, Buzzard IM, Gebhardt SE. Procedures for estimating nutrient values for food composition databases. J Food Comp and Anal. 1997;10:102–114.

36. Expert Panel on Detection, Evaluation, and Treatment of High Blood Cholesterol in Adults. Executive Summary of The Third Report of The National Cholesterol Education Program (NCEP) Expert Panel on Detection, Evaluation, And Treatment of High Blood Cholesterol In Adults (Adult Treatment Panel III). JAMA. 2001; 285(19):2486–2497. [PubMed: 11368702]

37. Clinical guidelines on the identification, evaluation, and treatment of overweight and obesity in adults: the evidence report [NIH Publication No. 98–4083]. National Heart, Lung, and Blood Institute; http://www.nhlbi.nih.gov/guidelines/obesity/ob_gdlns.pdf

38. Lean MEJ, Han TS, Morrison CE. Waist circumference as a measure for indicating need for weight management BMJ 1995; 311:158 doi:10.1136/bmj.311.6998.158

39. Nickerson BS. Evaluation of Obesity Cutoff Values in Hispanic Adults: Derivation of New Standards. J Clin Densitom. 2021;24(3):388–396. doi:10.1016/j.jocd.2020.10.010

40. American Diabetes Association; Standards of Medical Care in Diabetes—2010. Diabetes Care 1 January 2010; 33 (Supplement_1): S11–S61. https://doi.org/10.2337/dc10-S011

41. Chobanian AV, Bakris GL, Black HR, et al. National Heart, Lung, and Blood Institute Joint National Committee on Prevention, Detection, Evaluation, and Treatment of High Blood Pressure; National High Blood Pressure Education Program Coordinating Committee. The Seventh Report of the Joint National Committee on Prevention, Detection, Evaluation, and Treatment of High Blood Pressure: the JNC 7 report. JAMA. 2003; 289(19):2560–2572. [PubMed: 12748199]

42. D’Agostino RB Sr, Vasan RS, Pencina MJ, Wolf PA, Cobain M, Massaro JM, Kannel WB. General cardiovascular risk profile for use in primary care: the Framingham Heart Study. Circulation. 2008 Feb 12;117(6):743–53. doi: 10.1161/CIRCULATIONAHA.107.699579. Epub 2008 Jan 22. PMID: 18212285.

43. Castro V. (2021) CVrisk: Compute Risk Scores for Cardiovascular Diseases. https://cran.rstudio.com/web/packages/CVrisk/index.html. Accessed on 7/25/2022.

44. https://sites.cscc.unc.edu/hchs/study-data-and-analytic-methods-pub Hispanic Community Health Study/Study of Latinos (HCHS/SOL) Baseline Derived Variable Dictionary Version 4.23 (January 2020). HCHS/SOL Coordinating Center, Collaborative Studies Coordinating Center, Department of Biostatistics, UNC Chapel Hill. Accessed on 1/21/2023

45. Lumley T. (2020) “survey: analysis of complex survey samples”. R package version 4.0.

46. Lumley T. (2004) Analysis of complex survey samples. Journal of Statistical Software 9(1): 1–19

47. Cronbach, LJ. Coefficient alpha and the internal structure of tests. Psychometrika 1951; 18: 297–334

48. R Core Team (2020). R: language and environment for statistical computing. R Foundation for Statistical Computing, Vienna, Austria. URL https://www.R-project.org/.

49. Revelle W. (2021) psych: Procedures for Personality and Psychological Research, Northwestern University, Evanston, Illinois, USA, https://CRAN.R-project.org/package=psych Version = 2.1.9,.

50. Wickham H, Miller E. (2020). haven: Import and Export ‘SPSS’, ‘Stata’ and ‘SAS’ Files. R package version 2.3.1. https://CRAN.R-project.org/package=haven

51. Wickham H. (2016) ggplot2: Elegant Graphics for Data Analysis. Springer-Verlag New York, 2016.

52. Wickham, et al., (2019). Welcome to the tidyverse. Journal of Open Source Software, 4(43), 1686, https://doi.org/10.21105/joss.01686

53. Befort CA, Nazir N, Perri MG. (2012), Prevalence of Obesity Among Adults From Rural and Urban Areas of the United States: Findings From NHANES (2005-2008). The Journal of Rural Health, 28: 392–397. https://doi.org/10.1111/j.1748-0361.2012.00411.x

54. Kershaw KN, Giacinto RE, Gonzalez F, Isasi CR, Salgado H, Stamler J, Talavera GA, Tarraf W, Van Horn L, Wu D, Daviglus ML. Relationships of nativity and length of residence in the US with favorable cardiovascular health among Hispanics/Latinos: the Hispanic Community Health Study/Study of Latinos (HCHS/SOL). Preventive medicine. 2016 Aug 1;89:84–9.

55. Chen Y, Chen G, Abittan N, Xing J, Mossavar-Rahmani Y, Sotres-Alvarez D, Mattei J, Daviglus M, Isasi CR, Hu FB, Kaplan R, Qi Q, Healthy dietary patterns and risk of cardiovascular disease in US Hispanics/Latinos: the Hispanic Community Health Study/Study of Latinos (HCHS/SOL), The American Journal of Clinical Nutrition, 2022

56. Joyce BT, Wu D, Hou L, Dai Q, Castaneda SF, Gallo, LC, Talavera GA, Sotres-Alvarez, D, Van Horn L, Beasley J et al. DASH diet and prevalent metabolic syndrome in the Hispanic Community Health Study/Study of Latinos. Prev. Med. Rep. 2019, 15, 100950

57. Liu J, Huang Y, Dai Q, Fulda KG, Chen S, Tao M-H. Trends in Magnesium Intake among Hispanic Adults, the National Health and Nutrition Examination Survey (NHANES) 1999–2014. Nutrients. 2019; 11(12):2867. https://doi.org/10.3390/nu11122867

58. Jackson SL, King SMC, Zhao L, Cogswell ME. (2016). Prevalence of Excess Sodium Intake in the United States — NHANES, 2009–2012. Morbidity and Mortality Weekly Report, 64(52), 1393–1397. https://www.jstor.org/stable/24856953

59. Stephenson BJ, Wu SM, Dominici F. Identifying Dietary Consumption Patterns from Survey Data: A Bayesian Nonparametric Latent Class Model. medRxiv. 2022 Jan 1:2021–11.

60. Baranowski, T. 24-Hour Recall and Diet Record Methods, Nutritional Epidemiology, 3rd edn, Monographs in Epidemiology and Biostatistics (2012; online edn, Oxford Academic, 24 Jan. 2013)

61. Ravelli MN, Schoeller DA. Traditional Self-Reported Dietary Instruments Are Prone to Inaccuracies and New Approaches Are Needed. Front Nutr. 2020;7:90. doi:10.3389/fnut.2020.00090

62. Mossavar-Rahmani Y, Shaw PA, Wong WW, et al. Applying Recovery Biomarkers to Calibrate Self-Report Measures of Energy and Protein in the Hispanic Community Health Study/Study of Latinos. Am J Epidemiol. 2015;181(12):996–1007. doi:10.1093/aje/kwu468

