## Supplementary Data for "Examining Generalizability of Nutrient-Based Food Patterns and Their Cross-Sectional Associations with Cardiometabolic Health for Hispanic/Latino Adults in the US: Results from the National Health and Nutrition Examination Survey (NHANES) and the Hispanic Community Health Study/Study of Latinos (HCH"

**Supplementary Material**

**Cronbach’s coefficient alphas**

To assess reliability of the derived factors, we evaluated the internal consistency of those nutrients that load more than |0.40| on any factor using standardized Cronbach's coefficient alphas. For each factor, we calculated coefficient alphas for the overall nutrients and when item deleted for each nutrient loading.

For NHANES, standardized Cronbach's coefficient alphas for each factor were 0.966, 0.956, 0.922, 0.953, and 0.862, respectively. Most of the standardized Cronbach's coefficient alphas, when item deleted, were generally greater than 0.85 (results shown in Table 1 below).

Supplementary Table 1: Cronbach’s coefficient alphas for NHANES

| **Nutrient** | **Overall** | **Factor 1** | **Factor 2** | **Factor 3** | **Factor 4** | **Factor 5** |
| --- | --- | --- | --- | --- | --- | --- |
| Overall | 0.962 | 0.966 | 0.956 | 0.922 | 0.953 | 0.862 |
| Total Protein (gm) | 0.961 | 0.965 | 0.954 |  |  |  |
| Total Carbohydrate (gm) | 0.962 |  |  |  | 0.956 |  |
| Total Carbohydrate (gm) | 0.962 |  |  |  | 0.953 |  |
| Total Sugars (gm) | 0.963 |  |  |  |  |  |
| Total Dietary Fiber (gm) | 0.962 |  |  |  | 0.956 |  |
| Total Fat (gm) | 0.962 |  | 0.951 |  |  |  |
| Total MUFA (gm) | 0.962 |  | 0.952 |  |  |  |
| Total PUFA (gm) | 0.962 |  | 0.956 |  |  |  |
| Cholesterol (mg) | 0.963 | 0.970 | 0.961 |  |  |  |
| Vitamin E (mg) | 0.962 |  | 0.96 |  |  | 0.876 |
| Retinol (mcg) | 0.963 |  |  | 0.917 |  |  |
| Vitamin A (mcg) | 0.962 |  |  | 0.923 |  | 0.863 |
| Alpha-Carotene (mcg) | 0.964 |  |  |  |  | 0.868 |
| Beta-Carotene (mcg) | 0.964 |  |  |  |  | 0.842 |
| Beta-Cryptoxanthin (mcg) | 0.964 |  |  |  |  |  |
| Lycopene (mcg) | 0.964 |  |  |  |  |  |
| Lutein+Zeaxanthin (mcg) | 0.963 |  |  |  |  | 0.849 |
| Vitamin B1 (mg) | 0.962 | 0.966 |  |  |  |  |
| Vitamin B2 (mg) | 0.961 | 0.966 |  | 0.919 |  |  |
| Niacin (mg) | 0.962 | 0.966 |  |  |  |  |
| Vitamin B6 (mg) | 0.962 | 0.966 |  |  |  |  |
| Total Folate (mcg) | 0.962 | 0.967 |  |  | 0.955 |  |
| Vitamin B12 (mcg) | 0.962 | 0.968 |  | 0.926 |  |  |
| Vitamin C (mg) | 0.964 |  |  |  |  | 0.879 |
| Vitamin D (mcg) | 0.963 |  |  | 0.928 |  |  |
| Vitamin K (mcg) | 0.963 |  |  |  |  | 0.851 |
| Calcium (mg) | 0.962 |  |  | 0.917 |  |  |
| Phosphorus (mg) | 0.961 | 0.964 |  |  | 0.955 |  |
| Magnesium (mg) | 0.961 | 0.966 |  |  | 0.96 |  |
| Iron (mg) | 0.961 | 0.966 |  |  | 0.953 |  |
| Zinc (mg) | 0.962 | 0.966 | 0.957 |  |  |  |
| Copper (mg) | 0.961 | 0.966 |  |  | 0.952 |  |
| Sodium (mg) | 0.962 | 0.966 | 0.955 |  |  |  |
| Potassium (mg) | 0.961 | 0.966 |  |  | 0.954 |  |
| Selenium (mcg) | 0.962 | 0.965 | 0.955 |  |  |  |
| Caffeine (mg) | 0.965 |  |  |  |  |  |
| SFA4 (gm) | 0.963 |  |  | 0.921 |  |  |
| MCSFA (gm) | 0.963 |  |  | 0.923 |  |  |
| LCSFA (gm) | 0.962 |  | 0.954 | 0.926 |  |  |

For HCHS/SOL, standardized Cronbach's coefficient alphas for each factor were 0.961, 0.969, 0.923, 0.932, 0.884, 0.941, respectively. Most of the standardized Cronbach's coefficient alphas, when item deleted, were greater than 0.86 (results shown in Table 2 below).

Supplementary Table 2: Cronbach’s coefficient alphas for HCHS/SOL

| **Nutrient** | **Overall** | **Factor 1** | **Factor 2** | **Factor 3** | **Factor 4** | **Factor 5** |
| --- | --- | --- | --- | --- | --- | --- |
| Overall | 0.961 | 0.969 | 0.923 | 0.932 | 0.884 | 0.941 |
| Total Protein (gm) | 0.960 | 0.968 |  |  |  | 0.930 |
| Total Carbohydrate (gm) | 0.960 | 0.969 |  |  |  |  |
| Total Sugars (gm) | 0.962 |  |  |  |  |  |
| Total Dietary Fiber (gm) | 0.961 | 0.970 |  |  |  |  |
| Total Fat (gm) | 0.960 |  |  | 0.917 |  |  |
| Total MUFA (gm) | 0.961 |  |  | 0.921 |  |  |
| Total PUFA (gm) | 0.961 |  |  | 0.931 |  |  |
| Cholesterol (mg) | 0.961 |  |  | 0.945 |  | 0.954 |
| Vitamin E (mg) | 0.961 | 0.970 |  | 0.944 |  |  |
| Retinol (mcg) | 0.962 |  | 0.920 |  |  |  |
| Vitamin A (mcg) | 0.961 |  | 0.932 |  | 0.878 |  |
| Alpha-Carotene (mcg) | 0.963 |  |  |  | 0.888 |  |
| Beta-Carotene (mcg) | 0.962 |  |  |  | 0.868 |  |
| Beta-Cryptoxanthin (mcg) | 0.963 |  |  |  |  |  |
| Lycopene (mcg) | 0.963 |  |  |  |  |  |
| Lutein+Zeaxanthin (mcg) | 0.962 |  |  |  | 0.876 |  |
| Vitamin B1 (mg) | 0.960 | 0.968 |  |  |  |  |
| Vitamin B2 (mg) | 0.960 | 0.970 | 0.915 |  |  |  |
| Niacin (mg) | 0.960 | 0.969 |  |  |  | 0.945 |
| Vitamin B6 (mg) | 0.960 | 0.968 |  |  |  |  |
| Total Folate (mcg) | 0.960 | 0.969 |  |  |  |  |
| Vitamin B12 (mcg) | 0.961 |  | 0.923 |  |  |  |
| Vitamin C (mg) | 0.962 |  |  |  | 0.897 |  |
| Vitamin D (mcg) | 0.962 |  | 0.924 |  |  |  |
| Vitamin K (mcg) | 0.962 |  |  |  | 0.885 |  |
| Calcium (mg) | 0.961 |  | 0.916 |  |  |  |
| Phosphorus (mg) | 0.960 | 0.968 | 0.920 |  |  | 0.938 |
| Magnesium (mg) | 0.960 | 0.968 |  |  |  |  |
| Iron (mg) | 0.960 | 0.968 |  |  |  |  |
| Zinc (mg) | 0.960 | 0.968 |  |  |  | 0.940 |
| Copper (mg) | 0.960 | 0.969 |  |  |  |  |
| Sodium (mg) | 0.961 |  |  | 0.936 |  | 0.946 |
| Potassium (mg) | 0.960 | 0.968 |  |  | 0.892 |  |
| Selenium (mcg) | 0.960 | 0.969 |  |  |  | 0.934 |
| Caffeine (mg) | 0.964 |  |  |  |  |  |
| SFA4 (gm) | 0.962 |  | 0.924 |  |  |  |
| MCSFA (gm) | 0.962 |  | 0.926 |  |  |  |
| LCSFA (gm) | 0.961 |  | 0.926 | 0.926 |  |  |

**Coefficient of factor congruence**

The bolded congruence coefficient values are all greater than 0.90 indicating a high degree of factor similarity.

Supplementary Table 3: Congruence coefficients for NHANES and HCHS/SOL factors (F)

|  | HCHS/SOL meats  (F5) | HCHS/SOL fats/oils  (F3) | HCHS/SOL dairy  (F2) | HCHS/SOL grains/  legumes (F1) | HCHS/SOL fruits/  veggies (F4) |
| --- | --- | --- | --- | --- | --- |
| NHANES  meats (F1) | **0.95** | 0.66 | 0.67 | 0.88 | 0.44 |
| NHANES  fats/oils (F2) | 0.78 | **0.99** | 0.55 | 0.66 | 0.32 |
| NHANES  dairy (F3) | 0.55 | 0.54 | **0.98** | 0.57 | 0.38 |
| NHANES  grains/  legumes (F4) | 0.64 | 0.63 | 0.55 | **0.97** | 0.54 |
| NHANES  fruits/  veggies (F5) | 0.35 | 0.29 | 0.30 | 0.57 | **0.99** |

**Flow Diagrams**

Supplementary Figure 1: Flow diagrams for the identification of the analysis sample. Figure 1A: NHANES cohort flow diagram. Figure 1B: HCHS/SOL cohort flow diagram

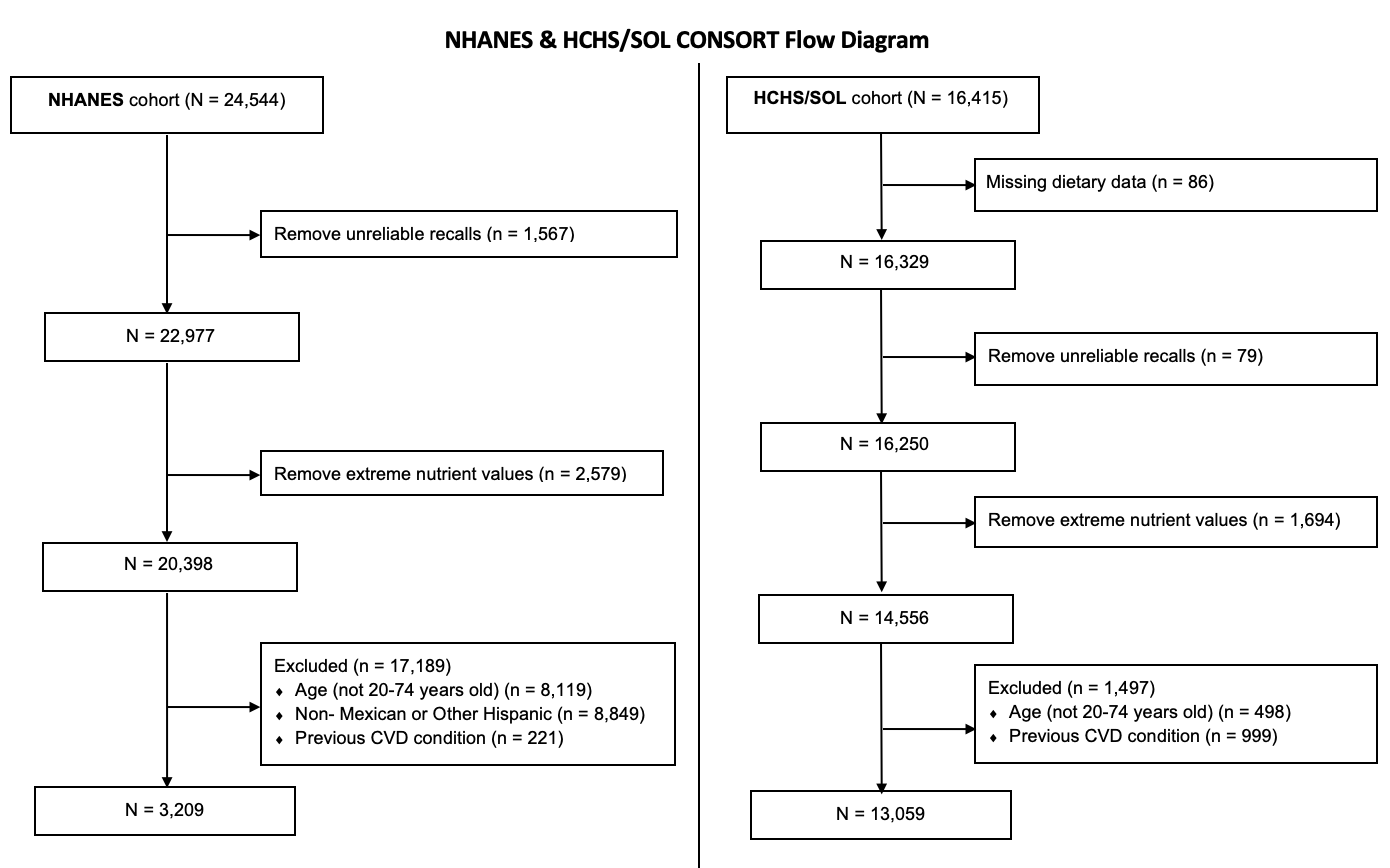

Note: Extreme nutrient values were defined as values below the 0.5th percentile and above the 99.5th percentile. Previous CVD conditions include heart failure, coronary heart disease, angina, heart attack, or stroke.

Supplementary Figure 2: Heatmap of odds ratio for cardiometabolic risk factors in single factor models. Stars indicate significance at p-value < 0.05, blue stars indicate negative associations while red stars reflect positive associations.

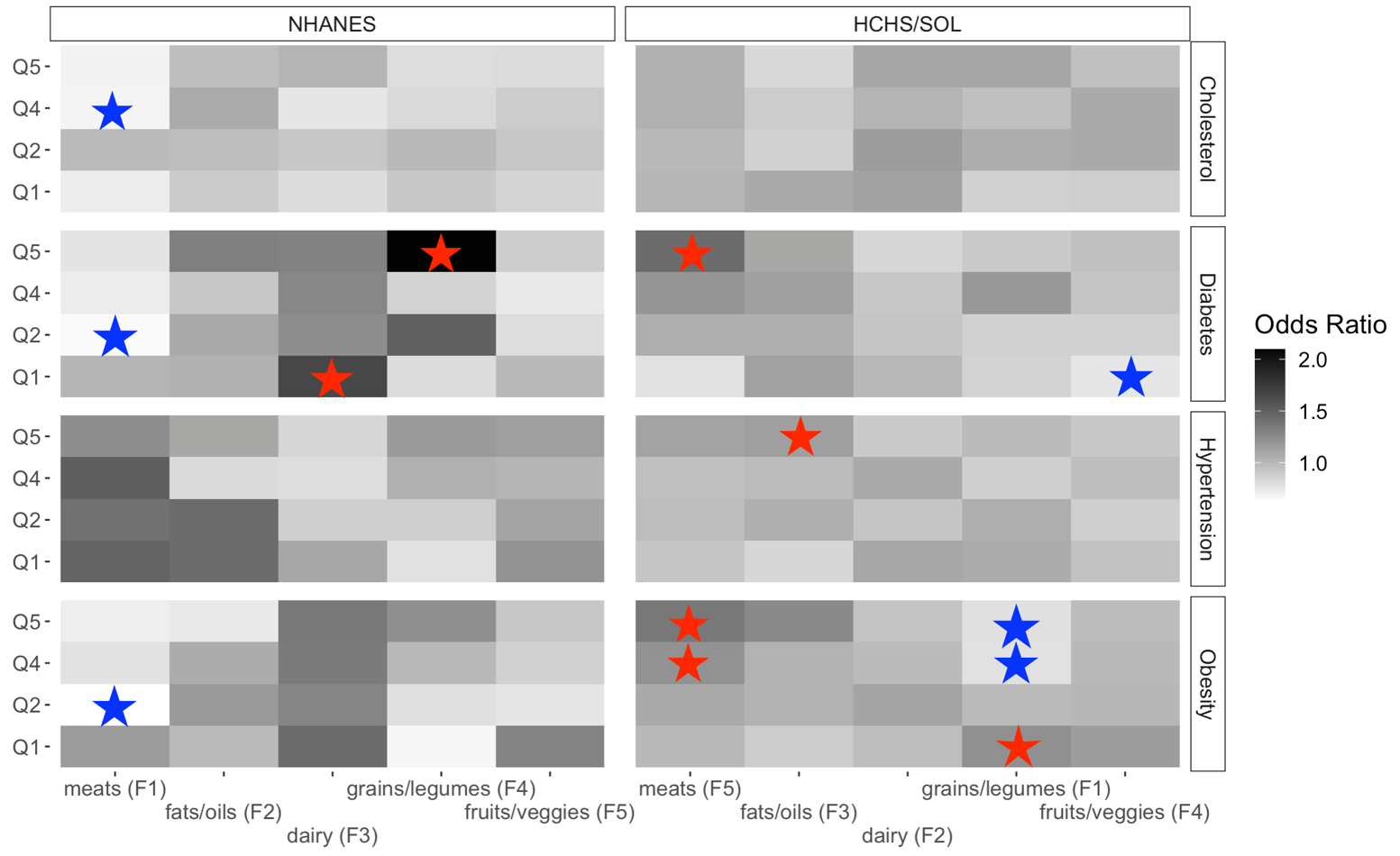
Note: Q, quintile; Conditions were defined for high cholesterol (total cholesterol ≥ 240 mg/dL, LDL cholesterol ≥ 160 mg/dL, HDL cholesterol < 40 mg/dL, self-reported use of cholesterol-lowering medication, or self-reported hypercholesterolemia), diabetes (fasting time > 8 hours & fasting plasma glucose ≥ 126 mg/dL, fasting time 8 hours and fasting glucose ≥ 200 mg/dL, or post-OGTT glucose ≥ 200 mg/dL, HbA1c ≥ 6.5%, self-reported medication use, or self-reported physician diagnosis), hypertension (BP ≥140/90 mm Hg or medication use), obesity [BMI ≥ 30 kg/m^2^ for 20-44 years old & waist circumference (women > 88 cm, men > 102 cm) for 45-74 years old], and hypertension (systolic blood pressure > 140 mm Hg, or diastolic blood pressure ≥ 90 mm Hg, or self-reported medication use).
